# Optimizing HPV Vaccination for Cervical Cancer Prevention in Women Living with HIV: A Cost-Effectiveness Analysis Across 13 African Countries

**DOI:** 10.64898/2026.10.05.26364501

**Authors:** Sahar Trabelsi, Kaja Abbas, Taylor A. Holroyd, Caroline L. Trotter, Amira Kebir, Slimane Ben Miled

## Abstract

**Background:** Women living with HIV face a substantially higher risk of persistent HPV infection and cervical cancer, yet HPV vaccination strategies targeting this high-risk population remain limited in many low- and middle-income countries. We evaluated the lifetime health impact and cost-effectiveness of HIV-targeted HPV vaccination strategies across 13 sub-Saharan African countries.

**Methods:** We developed an age-structured Markov model of HPV infection and cervical cancer in a closed cohort of 100,000 females, stratified by HIV status, and calibrated it separately for each country to age-specific cervical cancer incidence, cervical cancer mortality, and female HIV prevalence. Six HIV-targeted strategies were compared with routine two-dose vaccination at age 14 years. The strategies either provided a booster dose to previously vaccinated WLHIV or combined a booster for previously vaccinated WLHIV with a three-dose primary catch-up course for previously unvaccinated WLHIV. Interventions were delivered at ages 18 or 24 years under standard- and high-coverage assumptions. Outcomes included cervical cancer cases averted, cervical cancer deaths averted among WLHIV, discounted disability-adjusted life-years (DALYs) averted, additional physical vaccine doses, and incremental costs from the health-system and vaccination-programme perspectives.

**Results:** All HIV-targeted vaccination strategies reduced cervical cancer burden relative to routine vaccination across the 13 countries. Higher-coverage strategies generally produced greater health gains, with mean DALYs averted ranging from 9.05 for booster vaccination at age 24 years to 88.42 for high-coverage combined booster and catch-up vaccination at age 24 years. However, the relative performance of vaccination at ages 18 and 24 varied across strategies and countries. Booster-only strategies achieved the greatest health gains per additional vaccine dose, while combined strategies at higher coverage produced larger overall benefits. From the health-system perspective, 68 of 78 country–strategy comparisons were more effective and less costly than routine vaccination after accounting for avoided cervical cancer treatment costs. From the vaccination-programme perspective, all strategies required additional expenditure.

**Conclusions:** Targeted HPV vaccination among women living with HIV can reduce cervical cancer burden and may be cost-saving when avoided treatment costs are considered. Higher-coverage combined strategies produced the greatest overall health gains, while booster-only strategies were more dose-efficient, supporting context-specific vaccination decisions.

## 1. Introduction

Regions with a high burden of cervical cancer (CC) frequently overlap with settings where human immunodeficiency virus (HIV) incidence and prevalence are high. HIV infection increases susceptibility to human papillomavirus (HPV) acquisition and persistence through impaired immunity, leading to reduced HPV clearance and accelerated progression from infection to precancerous lesions and invasive CC [1]. As a consequence, women living with HIV experience substantially higher CC risk than women without HIV, with pooled estimates suggesting an approximately six-fold higher risk of cervical cancer [2]. In many low- and middle-income countries (LMICs), where both HPV vaccination and cervical cancer screening coverage remain low, CC incidence and mortality among women living with HIV remain high despite the expansion of antiretroviral therapy (ART) [3].

Prophylactic HPV vaccination is among the most effective interventions for CC prevention and is a cornerstone of the World Health Organization (WHO) global strategy for cervical cancer elimination [4]. Three HPV vaccine formulations are currently available globally: bivalent, quadrivalent, and nonavalent vaccines, all of which have demonstrated high efficacy in preventing vaccine-type HPV infections and related cervical lesions [5]. Evidence from immunogenicity and safety studies indicates that HPV vaccines are immunogenic and have acceptable safety profiles among people living with HIV [6]. WHO recommends at least two HPV vaccine doses and, where feasible, three doses for individuals known to be immunocompromised or living with HIV [4].

In this study, we assess the lifetime health impact and cost-effectiveness of extending HPV vaccination among women living with HIV beyond routine adolescent vaccination programmes across 13 African countries. Using an age-structured, lifetime cohort Markov model calibrated separately to country-specific cervical cancer incidence, cervical cancer mortality, and HIV prevalence, we compare the routine vaccination baseline (S0) with six HIV-targeted strategies delivered at ages 18 or 24 years. These include booster-only strategies for previously vaccinated women and combined strategies providing a booster dose to previously vaccinated women and a three-dose primary catch-up course to women who were not reached by routine vaccination programmes. Routine vaccination was evaluated at 50% coverage in all study countries and remained at this level under all strategies. The HIV-targeted components of S1–S4 were also evaluated at 50% coverage, whereas coverage of the HIV-targeted booster and catch-up components was increased to 98% under S5 and S6. Outcomes include cervical cancer cases averted, cervical cancer deaths averted among women living with HIV, disability-adjusted life-years (DALYs) averted, additional physical vaccine doses administered, and incremental discounted costs from the health-system and vaccination-programme perspectives. We also examine dose-normalized health and economic outcomes to compare marginal returns per additional vaccine dose and inform strategy selection in settings where vaccine supply or delivery capacity may be constrained.

We report incremental cost-effectiveness ratios (ICERs) for each strategy relative to the routine vaccination baseline (S0), together with sequential ICERs among non-dominated strategies on country-specific cost-effectiveness frontiers.

## 2. Methods

### 2.1 Model description

We developed an age-structured, annual-cycle state-transition (Markov) model to simulate oncogenic human papillomavirus (HPV) infection and cervical cancer over the lifetime of a closed cohort of 100,000 females, stratified by HIV status (Figure 1). Individuals enter the model at age 0 and are followed in single-year age groups up to age 100 years or until death. No births or immigration are introduced during the simulation. The model contains interconnected HIV-negative and HIV-positive strata to represent differences in HPV acquisition, clearance, precancer progression, cervical cancer recovery, and mortality by HIV status. HIV-negative individuals may acquire HIV during follow-up and move to the corresponding HIV-positive health state without resetting their HPV or cervical cancer history.

**Figure 1:**
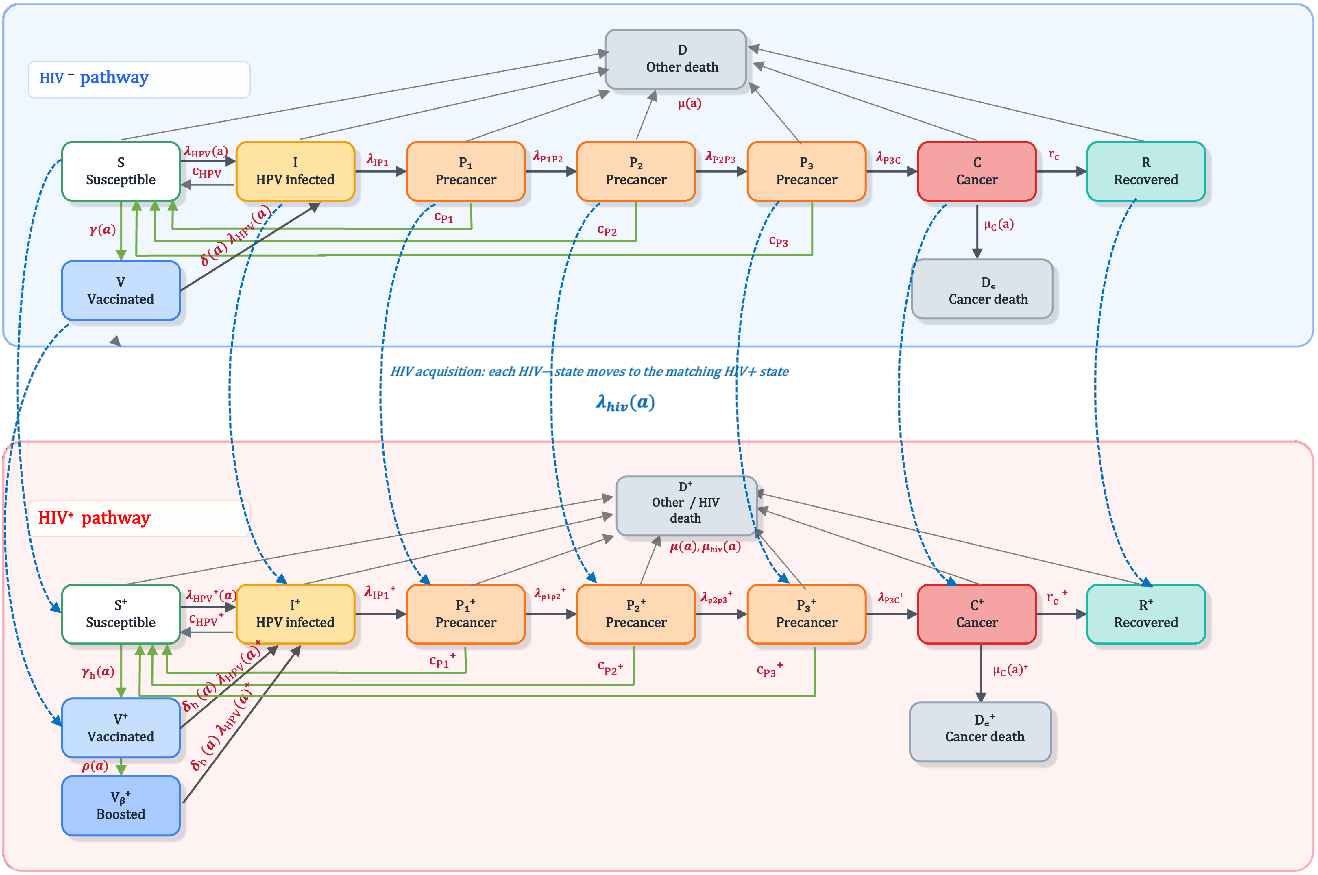
Schematic representation of the age-structured HPV–HIV state-transition model. The upper panel represents the HIV-negative pathway and the lower panel represents the HIV-positive pathway. Individuals progress through susceptible, vaccinated, HPV-infected, three sequential precancer, invasive cervical cancer, and post-treatment survivor states. The HIV-positive pathway additionally includes a boosted-vaccination state. Solid grey arrows represent disease progression, recovery, and mortality; green arrows represent vaccination, reduced HPV acquisition, and lesion clearance; and dashed blue arrows represent HIV acquisition with movement to the corresponding HIV-positive state while retaining the current HPV or cervical cancer stage. Background and HIV-related deaths are tracked separately in the model but may be combined visually in the schematic.

Persistent infection with oncogenic HPV may lead to cervical intraepithelial neoplasia (CIN), a spectrum of precancerous cervical lesions commonly classified clinically as CIN1, CIN2, and CIN3. Because country-specific data on individual CIN stages were limited, these clinical categories were not modelled separately. Instead, the precancerous phase was represented using three sequential tunnel states, P_1_, P_2_, and P_3_. These states capture the time spent in the precancerous phase and the gradual progression from persistent HPV infection to invasive cervical cancer; they should not be interpreted as direct representations of CIN1, CIN2, and CIN3. Individuals may clear a lesion from any precancer state and return to susceptibility, progress to the subsequent precancer state, or, from P_3_, progress to invasive cervical cancer.

The model also includes susceptible, vaccinated, HPV-infected, invasive cervical cancer, and post-treatment survivor states for both HIV-negative and HIV-positive individuals. An additional boosted-vaccination state is included for previously vaccinated women living with HIV who receive a booster dose. Individuals who recover from invasive cervical cancer enter the corresponding post-treatment survivor state and remain subject to background mortality and, where applicable, HIV-related mortality. Cancer recurrence and subsequent movement back into the HPV natural-history pathway are not modelled explicitly.

HPV acquisition is represented using separate age-specific hazards for HIV-negative and HIV-positive individuals. Infection may clear at HIV-specific rates, with individuals returning to the corresponding susceptible state. No durable natural immunity is assumed following HPV clearance, allowing individuals to become reinfected. Progression through the precancer states and to invasive cervical cancer is also HIV-specific, allowing for differences by HIV status in HPV acquisition, clearance, and precancer progression, with women living with HIV assumed to have lower clearance and faster precancer progression.

Vaccination is modelled as a prophylactic intervention that reduces the hazard of acquiring HPV. It does not alter HPV clearance or disease progression after infection has occurred. Primary vaccination is represented through the vaccinated susceptible states, while booster vaccination among previously vaccinated women living with HIV restores protection and resets its duration according to the booster assumptions. Full model structure, transitions, and parameter definitions are provided in Figure 1, Appendix 1 and Table A1.

### 2.2 Vaccination scenarios

To address whether a targeted, dose-sparing expansion of HPV vaccination to older adolescents and young women living with HIV could deliver high impact and good value for money, we evaluated across all 13 study countries a set of HIV-specific vaccination scenarios that vary by target age group, prior vaccination status, and vaccination coverage.

The baseline strategy (S0) represents routine two-dose HPV vaccination delivered to HIV-negative and HIV-positive girls at age 14 years, assuming 50% coverage across all 13 study countries to ensure comparability between settings (Table 1).

**Table 1.** HPV vaccination strategies evaluated across the 13 study countries. Strategy S0 represents routine HPV vaccination at age 14 years. Strategies S1–S6 target adolescents and young women living with HIV. S1 and S2 provide one booster dose to previously vaccinated individuals at ages 18 and 24 years, respectively. S3 and S4 combine booster vaccination for previously vaccinated individuals with a three-dose primary catch-up course for previously unvaccinated individuals. S5 and S6 apply the corresponding combined strategies at 98% coverage among the targeted population living with HIV.

| Strategy | Target age | HIV status | Prior vaccination status | Vaccination delivered | Intervention type | Coverage assumption |
|---|---|---|---|---|---|---|
| S0: Baseline | 14 | HIV− and HIV+ | Not applicable | Two-dose routine primary vaccination | Routine HPV vaccination | 50% routine vaccination coverage in all study countries |
| S1 | 18 | HIV+ | Previously vaccinated | One additional booster dose | Booster-only strategy | 50% coverage among eligible previously vaccinated individuals living with HIV |
| S2 | 24 | HIV+ | Previously vaccinated | One additional booster dose | Booster-only strategy | 50% coverage among eligible previously vaccinated individuals living with HIV |
| S3 | 18 | HIV+ | Previously vaccinated or previously unvaccinated | S1 booster plus a three-dose primary catch-up course for previously unvaccinated individuals | Booster plus catch-up vaccination | 50% coverage of the HIV-targeted booster and catch-up components |
| S4 | 24 | HIV+ | Previously vaccinated or previously unvaccinated | S2 booster plus a three-dose primary catch-up course for previously unvaccinated individuals | Booster plus catch-up vaccination | 50% coverage of the HIV-targeted booster and catch-up components |
| S5 | 18 | HIV+ | Previously vaccinated or previously unvaccinated | S1 booster plus a three-dose primary catch-up course for previously unvaccinated individuals | High-coverage booster plus catch-up vaccination | 98% coverage of the HIV-targeted booster and catch-up components; routine vaccination remains at 50% |
| S6 | 24 | HIV+ | Previously vaccinated or previously unvaccinated | S2 booster plus a three-dose primary catch-up course for previously unvaccinated individuals | High-coverage booster plus catch-up vaccination | 98% coverage of the HIV-targeted booster and catch-up components; routine vaccination remains at 50% |

*Notes: Routine primary vaccination under S0 consists of two physical vaccine doses. Primary catch-up vaccination under S3–S6 consists of three physical doses for previously unvaccinated women living with HIV. Booster vaccination consists of one additional physical dose for previously vaccinated women living with HIV.*

Against this baseline, we evaluated six targeted strategies designed for adolescents and young women living with HIV, reflecting three programmatically relevant dimensions: (i) age at delivery (late adolescence versus early adulthood), (ii) prior vaccination history (previously vaccinated through routine programmes versus unvaccinated), and (iii) vaccination coverage. Strategies S1 and S2 assess a minimal additional-dose approach by introducing a single booster dose among HIV-positive individuals who previously completed routine HPV vaccination. The booster is delivered at age 18 years (S1) or 24 years (S2), allowing direct comparison of boosting in late adolescence versus early adulthood.

Strategies S3 and S4 evaluate combined booster and primary catch-up vaccination among HIV-positive adolescents and young women. Previously vaccinated individuals receive a single booster dose, while those who were not reached by routine programmes receive a full three-dose primary catch-up course. Both components are delivered at age 18 years (S3) or 24 years (S4).

Strategies S5 and S6 represent high-coverage extensions of S3 and S4, respectively. These strategies combine booster vaccination for previously vaccinated individuals and primary catch-up vaccination for previously unvaccinated individuals within the same target age group, delivered at age 18 years (S5) or 24 years (S6).

For S1–S4, vaccination coverage was set at 50% of each eligible HIV-positive population. For S5 and S6, a high-coverage assumption of 98% was explored to represent intensified vaccine delivery among women living with HIV. Routine vaccination coverage remained at 50% across all strategies and study countries.

### 2.3 Country selection

We selected 13 African countries based on Gavi eligibility [7], variation in HIV-attributable cervical cancer burden, and the availability of epidemiological data required for model calibration. These included Cameroon, Côte d’Ivoire, Gambia, Kenya, Lesotho, Malawi, Mozambique, Rwanda, Tanzania, Togo, Uganda, Zambia, and Zimbabwe. The selected countries represent different epidemiological settings, allowing comparison of HIV-targeted HPV vaccination strategies across countries.

### 2.4 Country-specific calibration and model initialization

We calibrated the model separately for each country by minimizing a weighted objective function comparing model outputs with age-specific cervical cancer incidence, mortality, and female HIV prevalence (<u>Appendix 2</u>).

#### 2.4.1 Initial conditions

We simulated a closed cohort of 100,000 females entering at age 0 years and followed them annually through age 100 years, or until death. At birth, individuals were allocated to HIV-negative and HIV-positive susceptible states using a common initial HIV prevalence assumption of 0.236% across all countries. No individuals were initially assigned to HPV infection, precancer, or cervical cancer states.

#### 2.4.2 Assumptions on vaccine efficacy and duration of protection

Assumptions regarding HPV vaccine efficacy and duration of protection were informed by available evidence and international guidance (<u>Table A8</u>). Vaccine efficacy and waning were specified separately by HIV status. Given the limited evidence on the long-term clinical effectiveness of booster vaccination among women living with HIV, booster efficacy and duration of protection were treated as exploratory modelling assumptions.

### 2.5 Health outcomes and cost-effectiveness

Health outcomes were expressed in disability-adjusted life-years (DALYs), calculated as the sum of years of life lost (YLL) due to premature cervical cancer mortality and years lived with disability (YLD). YLL were derived using age-specific cervical cancer deaths and corresponding remaining life expectancy. YLD were estimated using a prevalence-based approach, applying a disability weight of 0.303 [8], to the person-years spent in the invasive cervical cancer compartments among HIV-negative and HIV-positive women. Disability associated with HPV infection, precancerous lesions, and HIV infection was not included, as the analysis focused on cervical cancer-related health outcomes. Future health outcomes were discounted at an annual rate of 3%. For each strategy, DALYs averted were estimated relative to the baseline strategy S0.

Total costs were calculated by combining vaccination programme costs (vaccine acquisition and delivery for routine, catch-up, and booster doses) and cervical cancer management costs. Cancer care costs were obtained by multiplying the country-specific unit cost per cervical cancer case by the model-projected number of incident cases. Future costs were discounted at an annual rate of 3%.

Cost-effectiveness was evaluated from two perspectives: a health-system perspective including vaccination and cervical cancer management costs, and a vaccination-programme perspective including vaccination costs only. Direct incremental cost-effectiveness ratios (ICERs) were calculated as incremental cost per DALY averted relative to S0. Strategies yielding greater health gains at lower total cost were classified as cost-saving. Sequential ICERs were calculated after excluding strictly and extendedly dominated strategies to identify the cost-effectiveness frontier. Full computational details and mathematical formulations are provided in Appendix 3.

#### 2.5.1 Cost inputs

The HPV vaccine acquisition cost was set at USD 4.50 per dose, based on WHO vaccine procurement price information [9]. An additional delivery cost of USD 1.50 per dose was assumed, giving a total cost of USD 6.00 per administered dose. This assumption was informed by published HPV vaccine delivery-cost estimates in low- and middle-income countries [10]. Routine vaccination consisted of two doses, primary catch-up vaccination of three doses, and booster vaccination of one dose. Cervical cancer management costs were estimated using a base-case proxy of USD 2,526 per incident case, based on the average cost reported among cervical cancer patients who had undergone screening at a Tanzanian hospital [11]. Given the lack of comparable country-specific estimates, costs for the remaining 12 countries were extrapolated using GDP per capita ratios relative to Tanzania, based on World Bank data [12]. Country-specific unit costs are presented in Table A9, and the calculation is detailed in Appendix 3.

Total vaccination costs were calculated from the number of physical vaccine doses administered, while cervical cancer management costs were calculated from incident cervical cancer cases. The vaccination-programme perspective included vaccination costs only, whereas the health-system perspective included both vaccination and cervical cancer management costs. All costs and health outcomes (YLL, YLD, and DALYs) were discounted at an annual rate of 3%.

#### 2.5.2 Analytic perspectives

The economic evaluation was conducted from two perspectives. The primary health-system perspective included vaccination costs and cervical cancer management costs, accounting for potential savings from cervical cancer cases averted. The secondary vaccination-programme perspective included vaccine acquisition and delivery costs only, reflecting the expenditure required to implement the vaccination strategies. Both perspectives used the same health outcomes.

Patient out-of-pocket costs, transportation, productivity losses, informal care, and the opportunity costs of existing healthcare resources were excluded. Therefore, the analysis did not adopt a societal perspective or assess the broader budgetary impact of vaccination.

### 2.6 Sensitivity analysis

We conducted a deterministic one-way sensitivity analysis (OWSA) for Mozambique to assess how uncertainty in calibrated epidemiological parameters affects model outcomes. Each of the 17 calibrated parameters was varied independently by ±20% around its calibrated value, using five equally spaced values, while all other parameters were held constant. The model was rerun under the baseline vaccination strategy (S0), and outcomes included total discounted DALYs, total discounted health-system costs, and total physical vaccine doses administered. Results were summarized using tornado diagrams showing the minimum and maximum changes in each outcome and local sensitivity measures.

In addition, we performed a probabilistic sensitivity analysis (PSA) to jointly propagate uncertainty across the 17 calibrated epidemiological parameters. For each of 2,000 iterations, all parameters were simultaneously sampled from independent uniform distributions defined as ±10% around their calibrated values, and the model was run for each vaccination strategy. PSA outputs were converted into incremental costs and discounted DALYs averted relative to the baseline scenario (S0) and summarized using cost-effectiveness planes. Detailed descriptions of the calculation methods are provided in Appendix 4.

## 3. Results

This section reports the projected health impact and cost-effectiveness of HIV-targeted HPV vaccination strategies across 13 African countries. We compare booster vaccination among previously vaccinated women living with HIV with strategies combining booster and primary catch-up vaccination for those missed by routine programmes, under alternative assumptions on vaccination age and coverage. For each scenario, we estimate cervical cancer cases averted across both HIV strata, cervical cancer deaths averted among women living with HIV, discounted DALYs averted, and changes in vaccination programme and cervical cancer management costs. We first summarize the baseline epidemiological context, then present comparative health and economic outcomes across strategies and countries, and finally report sensitivity analyses for key epidemiological and economic parameters.

### 3.1 Baseline burden and vaccination context

Baseline cervical cancer incidence and mortality varied across the 13 study countries, as shown by the country-specific calibration data presented in Figure A1. In the model, country-specific calibrated epidemiology influenced the number of women eligible for additional vaccination and the magnitude of the projected health impact. For comparability across countries, a common baseline HPV vaccination coverage assumption of 50% was applied in the base-case analysis.

### 3.2 Health impact of vaccination strategies

All six vaccination strategies generated health gains relative to S0 across the 13 countries, although the magnitude varied substantially between settings (Table 2). Booster vaccination at age 18 (S1) averted a mean of 11.92 DALYs (SD: 11.80; range: 0.82–37.07), compared with 9.05 DALYs (SD: 11.69; range: 0.19–35.63) for booster vaccination at age 24 (S2). The corresponding mean numbers of cervical cancer deaths averted among women living with HIV were 1.84 and 1.78, respectively. S1 generally produced greater health gains than S2, although the age-24 strategy generated slightly greater benefits in Mozambique and Zimbabwe.

**Table 2.** Multi-country health impact and marginal dose efficiency of HIV-targeted HPV vaccination strategies relative to baseline (S0) across 13 African countries.

| **Outcome** | **Cameroon** | **Côte d’Ivoire** | **Gambia** | **Kenya** | **Lesotho** | **Malawi** | **Mozambique** | **Rwanda** | **Tanzania** | **Togo** | **Uganda** | **Zambia** | **Zimbabwe** |
|---|---|---|---|---|---|---|---|---|---|---|---|---|---|
| **S1. Baseline + booster at age 18 (HIV+)** |  |  |  |  |  |  |  |  |  |  |  |  |  |
| **DALYs averted vs S0** | 1.30 | 3.00 | 0.82 | 15.91 | 27.38 | 13.55 | 8.82 | 4.12 | 25.48 | 1.56 | 13.99 | 37.07 | 1.99 |
| **HIV+ deaths averted vs S0** | 0.05 | 0.47 | 0.03 | 2.21 | 5.56 | 1.68 | 1.48 | 0.47 | 3.28 | 0.21 | 2.19 | 5.83 | 0.45 |
| **DALYs reduction (%) vs S0** | 0.02 | 0.05 | 0.02 | 0.17 | 0.25 | 0.06 | 0.12 | 0.03 | 0.19 | 0.02 | 0.11 | 0.21 | 0.04 |
| **Additional physical vaccine doses vs S0** | 56.8 | 46.3 | 34.6 | 49.8 | 794.3 | 39.4 | 513.7 | 10.0 | 151.9 | 18.9 | 84.8 | 209.2 | 109.6 |
| **DALYs averted per 100k doses** | 2,294.2 | 6,470.3 | 2,364.7 | 31,976.8 | 3,446.3 | 34,358.5 | 1,717.7 | 41,100.9 | 16,774.0 | 8,261.7 | 16,496.7 | 17,719.3 | 1,820.6 |
| **HIV+ deaths averted per 100k doses** | 81.6 | 1,011.4 | 83.0 | 4,432.6 | 700.4 | 4,268.8 | 287.8 | 4,639.2 | 2,159.2 | 1,128.2 | 2,577.6 | 2,788.9 | 410.3 |
| **S2. Baseline + booster at age 24 (HIV+)** |  |  |  |  |  |  |  |  |  |  |  |  |  |
| **DALYs averted vs S0** | 1.05 | 1.39 | 0.19 | 2.39 | 22.60 | 1.83 | 15.55 | 0.22 | 23.20 | 0.27 | 11.30 | 35.63 | 2.07 |
| **HIV+ deaths averted vs S0** | 0.08 | 0.26 | 0.01 | 0.40 | 5.66 | 0.30 | 3.06 | 0.03 | 3.68 | 0.05 | 2.11 | 6.95 | 0.57 |
| **DALYs reduction (%) vs S0** | 0.02 | 0.02 | 0.00 | 0.03 | 0.21 | 0.01 | 0.21 | 0.00 | 0.18 | 0.00 | 0.09 | 0.20 | 0.04 |
| **Additional physical vaccine doses vs S0** | 49.4 | 17.4 | 12.2 | 15.3 | 607.2 | 5.0 | 1,030.6 | 0.5 | 122.7 | 9.0 | 87.1 | 186.7 | 120.0 |
| **DALYs averted per 100k doses** | 2,115.1 | 8,010.4 | 1,575.3 | 15,647.6 | 3,722.1 | 36,342.1 | 1,509.2 | 45,124.8 | 18,912.0 | 2,947.7 | 12,972.8 | 19,083.9 | 1,721.9 |
| **HIV+ deaths averted per 100k doses** | 162.0 | 1,494.4 | 104.9 | 2,626.7 | 931.4 | 5,992.1 | 297.3 | 6,738.8 | 2,996.8 | 503.6 | 2,420.6 | 3,723.7 | 472.2 |
| **S3. S1 + three-dose catch-up primary at age 18 (HIV+), standard coverage** |  |  |  |  |  |  |  |  |  |  |  |  |  |
| **DALYs averted vs S0** | 4.08 | 7.03 | 2.39 | 49.28 | 72.55 | 50.18 | 27.05 | 12.75 | 79.12 | 4.84 | 43.01 | 101.40 | 5.85 |
| **HIV+ deaths averted vs S0** | 0.10 | 1.00 | 0.07 | 6.47 | 13.23 | 5.79 | 4.13 | 1.34 | 9.78 | 0.63 | 6.02 | 14.54 | 1.16 |
| **DALYs reduction (%) vs S0** | 0.06 | 0.11 | 0.05 | 0.53 | 0.67 | 0.21 | 0.36 | 0.11 | 0.60 | 0.06 | 0.34 | 0.58 | 0.11 |
| **Additional physical vaccine doses vs S0** | 337.9 | 291.2 | 206.6 | 330.8 | 5,222.1 | 377.2 | 3,109.1 | 81.7 | 700.4 | 109.9 | 696.8 | 1,162.2 | 903.0 |
| **DALYs averted per 100k doses** | 1,206.0 | 2,415.4 | 1,154.7 | 14,895.4 | 1,389.3 | 13,305.3 | 869.9 | 15,592.5 | 11,295.7 | 4,401.9 | 6,172.4 | 8,724.6 | 647.9 |
| **HIV+ deaths averted per 100k doses** | 29.4 | 344.6 | 31.9 | 1,956.8 | 253.3 | 1,534.7 | 132.7 | 1,636.5 | 1,397.0 | 570.5 | 863.7 | 1,250.8 | 128.3 |
| **S4. S2 + three-dose catch-up primary at age 24 (HIV+), standard coverage** |  |  |  |  |  |  |  |  |  |  |  |  |  |
| **DALYs averted vs S0** | 5.02 | 9.67 | 2.34 | 45.23 | 92.21 | 63.56 | 49.36 | 12.49 | 95.52 | 3.64 | 74.66 | 146.81 | 11.84 |
| **HIV+ deaths averted vs S0** | 0.27 | 1.69 | 0.14 | 7.47 | 20.44 | 9.36 | 8.96 | 1.64 | 14.36 | 0.61 | 12.63 | 25.36 | 2.83 |
| **DALYs reduction (%) vs S0** | 0.07 | 0.15 | 0.05 | 0.49 | 0.85 | 0.27 | 0.66 | 0.10 | 0.73 | 0.05 | 0.59 | 0.84 | 0.23 |
| **Additional physical vaccine doses vs S0** | 845.2 | 486.5 | 499.4 | 990.5 | 10,208.2 | 1,001.2 | 9,819.7 | 168.3 | 1,334.4 | 388.0 | 2,465.6 | 3,075.0 | 3,334.1 |
| **DALYs averted per 100k doses** | 594.2 | 1,986.8 | 469.5 | 4,566.9 | 903.3 | 6,348.2 | 502.6 | 7,424.6 | 7,158.9 | 938.5 | 3,028.1 | 4,774.5 | 355.2 |
| **HIV+ deaths averted per 100k doses** | 31.6 | 347.9 | 28.8 | 754.3 | 200.3 | 934.7 | 91.3 | 975.9 | 1,075.8 | 158.0 | 512.2 | 824.8 | 85.0 |
| **S5. S3 with high HIV+ coverage (98%)** |  |  |  |  |  |  |  |  |  |  |  |  |  |
| **DALYs averted vs S0** | 6.43 | 11.52 | 3.93 | 84.07 | 118.99 | 91.53 | 41.48 | 23.20 | 116.48 | 8.02 | 73.41 | 157.84 | 9.89 |
| **HIV+ deaths averted vs S0** | 0.16 | 1.65 | 0.11 | 10.99 | 21.44 | 10.43 | 6.33 | 2.40 | 14.18 | 1.03 | 10.26 | 22.26 | 1.96 |
| **DALYs reduction (%) vs S0** | 0.10 | 0.18 | 0.08 | 0.91 | 1.09 | 0.39 | 0.56 | 0.19 | 0.88 | 0.11 | 0.58 | 0.91 | 0.19 |
| **Additional physical vaccine doses vs S0** | 703.9 | 584.4 | 436.9 | 709.6 | 10,571.7 | 853.2 | 6,189.4 | 179.4 | 1,465.7 | 233.2 | 1,388.6 | 2,360.7 | 1,808.0 |
| **DALYs averted per 100k doses** | 913.8 | 1,970.6 | 899.0 | 11,847.1 | 1,125.5 | 10,728.0 | 670.3 | 12,932.2 | 7,947.2 | 3,438.7 | 5,286.9 | 6,686.0 | 547.1 |
| **HIV+ deaths averted per 100k doses** | 22.1 | 281.7 | 24.6 | 1,548.4 | 202.8 | 1,222.5 | 102.2 | 1,339.2 | 967.3 | 442.4 | 738.8 | 942.8 | 108.1 |
| **S6. S4 with high HIV+ coverage (98%)** |  |  |  |  |  |  |  |  |  |  |  |  |  |
| **DALYs averted vs S0** | 9.07 | 19.15 | 4.82 | 96.59 | 165.17 | 151.16 | 80.93 | 30.27 | 159.43 | 7.50 | 141.07 | 261.98 | 22.30 |
| **HIV+ deaths averted vs S0** | 0.47 | 3.33 | 0.29 | 15.87 | 36.13 | 22.09 | 14.73 | 3.94 | 23.63 | 1.26 | 23.74 | 44.43 | 5.30 |
| **DALYs reduction (%) vs S0** | 0.13 | 0.30 | 0.09 | 1.04 | 1.52 | 0.64 | 1.09 | 0.25 | 1.21 | 0.10 | 1.12 | 1.51 | 0.44 |
| **Additional physical vaccine doses vs S0** | 1,770.3 | 1,051.2 | 1,063.6 | 2,062.0 | 20,867.5 | 2,325.0 | 19,524.9 | 393.1 | 2,936.1 | 788.6 | 4,916.2 | 6,313.0 | 6,685.4 |
| **DALYs averted per 100k doses** | 512.3 | 1,822.1 | 453.1 | 4,684.4 | 791.5 | 6,501.5 | 414.5 | 7,700.8 | 5,429.9 | 951.6 | 2,869.4 | 4,149.8 | 333.5 |
| **HIV+ deaths averted per 100k doses** | 26.8 | 317.1 | 27.3 | 769.5 | 173.2 | 949.9 | 75.4 | 1,003.2 | 804.7 | 159.3 | 482.8 | 703.7 | 79.2 |

*Note:* DALY reduction is expressed as a percentage relative to S0. Dose-normalized outcomes are reported per 100,000 additional physical vaccine doses. Deaths averted refer to cervical cancer deaths among women living with HIV.

Strategies combining booster and primary catch-up vaccination generally produced larger absolute health gains than booster-only strategies. At age 18, mean DALYs averted increased from 35.35 (SD: 33.39; range: 2.39–101.40) under standard coverage (S3) to 57.45 (SD: 52.59; range: 3.93–157.84) under high coverage (S5). At age 24, the corresponding means increased from 47.11 (SD: 45.52; range: 2.34–146.81) under S4 to 88.42 (SD: 81.79; range: 4.82–261.98) under S6. Increasing coverage of the HIV-targeted components from 50% to 98% therefore strengthened the projected health impact of both combined strategies. For example, DALYs averted increased from 49.36 under S4 to 80.93 under S6 in Mozambique, from 74.66 to 141.07 in Uganda, and from 146.81 to 261.98 in Zambia.

S6 produced the largest average absolute health gain across countries, averting a mean of 15.02 cervical cancer deaths among women living with HIV. However, substantial cross-country variation was observed. Under S6, DALYs averted ranged from 4.82 in Gambia to 261.98 in Zambia. Relatively large gains were also observed in Lesotho (165.17), Tanzania (159.43), Malawi (151.16), and Uganda (141.07), compared with smaller gains in Cameroon (9.07), Togo (7.50), and Côte d’Ivoire (19.15). The pattern was also reflected in cervical cancer deaths averted among women living with HIV, ranging from 0.29 in Gambia to 44.43 in Zambia. Although S6 generated the greatest absolute DALY benefit in most countries, S5 produced slightly greater gains in Togo (8.02 versus 7.50 DALYs).

The age-specific profiles in Figure A2 show that annual cervical cancer cases among women living with HIV were concentrated mainly in middle adulthood, while cumulative cervical cancer deaths increased over the lifetime horizon. The magnitude and timing of these outcomes varied across the 13 countries, reflecting differences in the modelled cervical cancer burden. Differences between vaccination strategies were generally modest in the annual curves, but accumulated over time, resulting in reductions in cervical cancer cases and deaths.

Absolute health impact differed from marginal dose efficiency. Booster-only strategies required fewer additional physical vaccine doses and generated the highest average dose-normalized benefits. The arithmetic mean numbers of DALYs averted per 100,000 additional doses were 14,215.5 for S1 and 13,052.5 for S2, compared with 6,313.1 for S3, 3,003.9 for S4, 4,999.4 for S5, and 2,816.5 for S6. Thus, although S6 generated the largest average absolute health benefit, its benefit per additional dose was lower because it required substantially more vaccine doses.

Figure 2(a) further illustrates variation in dose efficiency across countries. The dose-weighted mean DALYs averted per 100,000 additional vaccine doses were 7,313.3, 5,200.2, 3,396.4, 1,769.0, 2,717.1, and 1,625.9 for S1–S6, respectively. These values were lower than the corresponding arithmetic means, indicating that countries contributing more additional doses generally had lower dose-normalized health returns. Given the substantial variation across countries, dose-normalized outcomes should be interpreted alongside absolute DALYs, cervical cancer cases and deaths averted, and the total number of vaccine doses administered.

**Figure 2.**
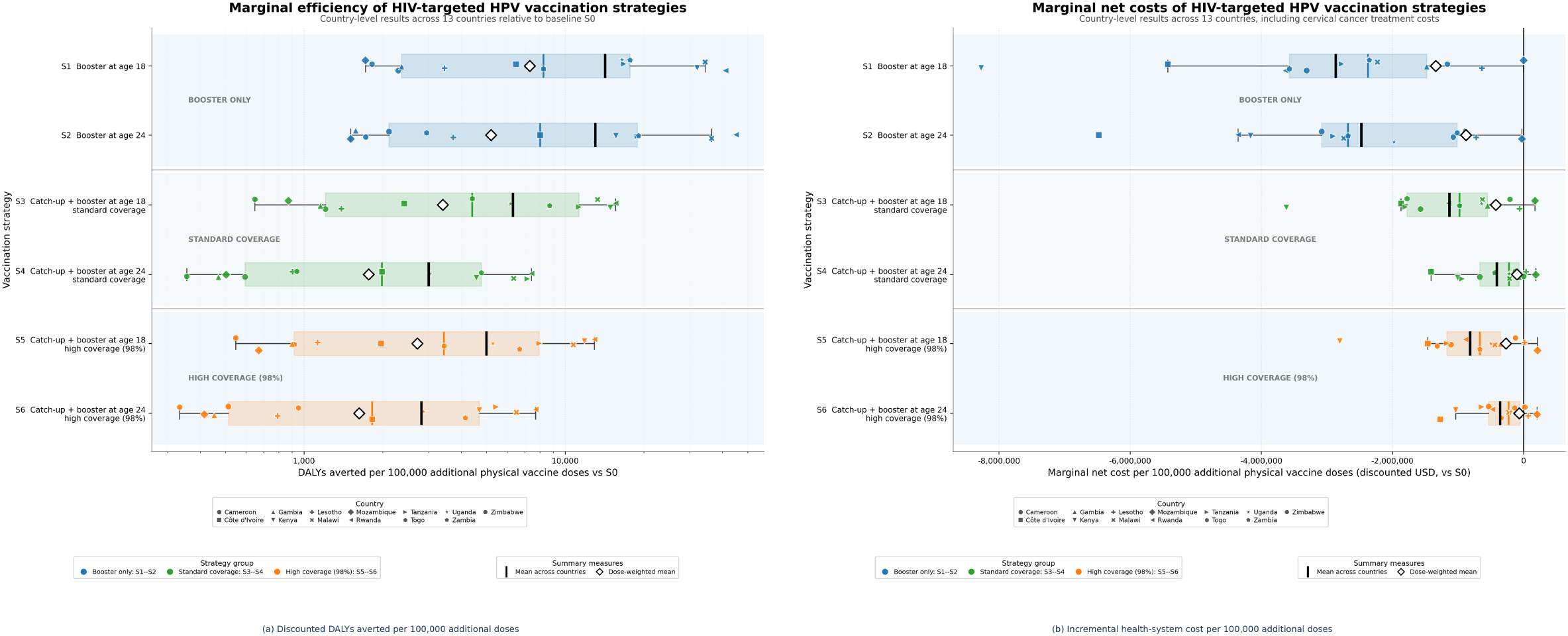
Marginal health and economic outcomes of HIV-targeted HPV vaccination strategies across 13 African countries. Outcomes are expressed per 100,000 additional physical vaccine doses relative to the baseline strategy (S0). Additional doses include booster and primary catch-up vaccination. Marker shapes identify countries, while colors distinguish vaccination strategies. Boxplots show the cross-country distributions, black markers indicate arithmetic means, and open diamonds represent dose-weighted means.

### 3.3 Economic outcomes and cost-effectiveness

Economic outcomes varied across countries and vaccination strategies, reflecting differences in vaccination expenditure and avoided cervical cancer management costs. Figure 3 presents the incremental discounted costs and DALYs averted under the health-system and vaccination-programme perspectives, while Tables 3 and 4 report the corresponding country-specific cost outcomes and direct ICERs.

**Table 3.** Discounted costs and cost-effectiveness of HIV-targeted HPV vaccination strategies relative to baseline (S0) across 13 African countries, including cervical cancer management costs.

| **Outcome** | **Cameroon** | **Côte d’Ivoire** | **Gambia** | **Kenya** | **Lesotho** | **Malawi** | **Mozambique** | **Rwanda** | **Tanzania** | **Togo** | **Uganda** | **Zambia** | **Zimbabwe** |
|---|---|---|---|---|---|---|---|---|---|---|---|---|---|
| **S0. Baseline routine vaccination** |  |  |  |  |  |  |  |  |  |  |  |  |  |
| **Discounted total cost (USD)** | 3,110,769 | 4,190,593 | 1,083,079 | 4,883,743 | 2,693,553 | 3,394,217 | 1,095,184 | 3,662,473 | 4,095,080 | 2,288,050 | 3,531,727 | 5,045,491 | 3,146,928 |
| **S1. Baseline + booster at age 18 (HIV+)** |  |  |  |  |  |  |  |  |  |  |  |  |  |
| **Discounted total cost (USD)** | 3,108,888 | 4,188,081 | 1,082,569 | 4,879,627 | 2,688,493 | 3,393,337 | 1,095,186 | 3,662,108 | 4,090,864 | 2,287,374 | 3,529,716 | 5,040,569 | 3,145,656 |
| **Incremental cost vs S0 (USD)** | -1,881 | -2,513 | -511 | -4,117 | -5,060 | -880 | 2 | -364 | -4,216 | -676 | -2,012 | -4,921 | -1,273 |
| **ICER vs S0 (USD/DALY averted)** | -1,442.17 | -838.50 | -625.00 | -258.74 | -184.82 | -64.91 | 0.22 | -88.38 | -165.51 | -432.42 | -143.78 | -132.76 | -637.94 |
| **S2. Baseline + booster at age 24 (HIV+)** |  |  |  |  |  |  |  |  |  |  |  |  |  |
| **Discounted total cost (USD)** | 3,109,246 | 4,189,466 | 1,082,969 | 4,883,108 | 2,689,158 | 3,394,079 | 1,094,902 | 3,662,451 | 4,091,515 | 2,287,959 | 3,530,006 | 5,040,491 | 3,145,638 |
| **Incremental cost vs S0 (USD)** | -1,522 | -1,127 | -110 | -636 | -4,395 | -138 | -282 | -22 | -3,565 | -92 | -1,722 | -4,999 | -1,290 |
| **ICER vs S0 (USD/DALY averted)** | -1,455.81 | -808.85 | -574.75 | -266.24 | -194.44 | -75.55 | -18.10 | -96.52 | -153.68 | -344.27 | -152.33 | -140.30 | -624.27 |
| **S3. S1 + three-dose catch-up primary at age 18 (HIV+), standard coverage** |  |  |  |  |  |  |  |  |  |  |  |  |  |
| **Discounted total cost (USD)** | 3,105,450 | 4,185,146 | 1,081,944 | 4,871,773 | 2,690,293 | 3,391,839 | 1,100,556 | 3,661,536 | 4,082,456 | 2,286,098 | 3,527,253 | 5,034,129 | 3,145,042 |
| **Incremental cost vs S0 (USD)** | -5,318 | -5,447 | -1,135 | -11,970 | -3,259 | -2,378 | 5,372 | -937 | -12,624 | -1,952 | -4,475 | -11,362 | -1,886 |
| **ICER vs S0 (USD/DALY averted)** | -1,304.97 | -774.33 | -475.73 | -242.92 | -44.92 | -47.39 | 198.63 | -73.53 | -159.56 | -403.66 | -104.04 | -112.05 | -322.43 |
| **S4. S2 + three-dose catch-up primary at age 24 (HIV+), standard coverage** |  |  |  |  |  |  |  |  |  |  |  |  |  |
| **Discounted total cost (USD)** | 3,105,136 | 4,183,730 | 1,082,729 | 4,873,782 | 2,697,434 | 3,392,043 | 1,113,366 | 3,661,711 | 4,082,562 | 2,287,549 | 3,526,270 | 5,031,972 | 3,146,971 |
| **Incremental cost vs S0 (USD)** | -5,633 | -6,863 | -350 | -9,962 | 3,881 | -2,174 | 18,182 | -762 | -12,518 | -501 | -5,458 | -13,518 | 42 |
| **ICER vs S0 (USD/DALY averted)** | -1,121.57 | -710.06 | -149.22 | -220.23 | 42.09 | -34.21 | 368.38 | -60.99 | -131.04 | -137.71 | -73.10 | -92.08 | 3.58 |
| **S5. S3 with high HIV+ coverage (98%)** |  |  |  |  |  |  |  |  |  |  |  |  |  |
| **Discounted total cost (USD)** | 3,102,963 | 4,182,041 | 1,081,542 | 4,863,842 | 2,695,042 | 3,390,469 | 1,108,277 | 3,660,877 | 4,077,902 | 2,284,977 | 3,524,770 | 5,029,738 | 3,144,691 |
| **Incremental cost vs S0 (USD)** | -7,805 | -8,552 | -1,537 | -19,902 | 1,489 | -3,748 | 13,094 | -1,596 | -17,178 | -3,073 | -6,958 | -15,753 | -2,237 |
| **ICER vs S0 (USD/DALY averted)** | -1,213.42 | -742.59 | -391.30 | -236.73 | 12.52 | -40.94 | 315.62 | -68.79 | -147.47 | -383.15 | -94.78 | -99.80 | -226.12 |
| **S6. S4 with high HIV+ coverage (98%)** |  |  |  |  |  |  |  |  |  |  |  |  |  |
| **Discounted total cost (USD)** | 3,101,296 | 4,177,219 | 1,082,463 | 4,862,369 | 2,707,884 | 3,388,922 | 1,134,793 | 3,660,591 | 4,076,198 | 2,286,980 | 3,522,196 | 5,023,870 | 3,148,160 |
| **Incremental cost vs S0 (USD)** | -9,473 | -13,375 | -616 | -21,375 | 14,332 | -5,295 | 39,609 | -1,882 | -18,882 | -1,070 | -9,532 | -21,621 | 1,232 |
| **ICER vs S0 (USD/DALY averted)** | -1,044.51 | -698.31 | -127.86 | -221.29 | 86.77 | -35.03 | 489.44 | -62.16 | -118.44 | -142.62 | -67.57 | -82.53 | 55.26 |

Note: Total costs include discounted vaccination programme and country-specific cervical cancer management costs. Incremental costs and ICERs are calculated relative to S0. ICERs are expressed in USD per discounted DALY averted. Negative ICERs indicate cost-saving strategies with positive health gains, whereas positive ICERs indicate additional expenditure per DALY averted. Small differences between total and incremental costs may arise from rounding.

**Table 4.** Discounted costs and cost-effectiveness of HIV-targeted HPV vaccination strategies relative to baseline (S0) across 13 African countries, excluding cervical cancer management costs.

| **Outcome** | **Cameroon** | **Côte d’Ivoire** | **Gambia** | **Kenya** | **Lesotho** | **Malawi** | **Mozambique** | **Rwanda** | **Tanzania** | **Togo** | **Uganda** | **Zambia** | **Zimbabwe** |
|---|---|---|---|---|---|---|---|---|---|---|---|---|---|
| **S0. Baseline routine vaccination** |  |  |  |  |  |  |  |  |  |  |  |  |  |
| **Discounted total cost (USD)** | 157,439 | 87,917 | 92,759 | 76,636 | 131,217 | 65,062 | 176,369 | 43,037 | 252,448 | 79,892 | 70,427 | 162,545 | 81,092 |
| **S1. Baseline + booster at age 18 (HIV+)** |  |  |  |  |  |  |  |  |  |  |  |  |  |
| **Discounted total cost (USD)** | 157,634 | 88,075 | 92,877 | 76,806 | 133,935 | 65,197 | 178,127 | 43,072 | 252,968 | 79,957 | 70,718 | 163,261 | 81,467 |
| **Incremental cost vs S0 (USD)** | 194 | 158 | 118 | 170 | 2,718 | 135 | 1,758 | 34 | 520 | 65 | 290 | 716 | 375 |
| **ICER vs S0 (USD/DALY averted)** | 149.14 | 52.88 | 144.70 | 10.70 | 99.29 | 9.96 | 199.20 | 8.33 | 20.40 | 41.42 | 20.74 | 19.31 | 187.94 |
| **S2. Baseline + booster at age 24 (HIV+)** |  |  |  |  |  |  |  |  |  |  |  |  |  |
| **Discounted total cost (USD)** | 157,581 | 87,967 | 92,794 | 76,680 | 132,957 | 65,076 | 179,322 | 43,039 | 252,799 | 79,918 | 70,677 | 163,080 | 81,436 |
| **Incremental cost vs S0 (USD)** | 142 | 50 | 35 | 44 | 1,740 | 14 | 2,953 | 1 | 352 | 26 | 250 | 535 | 344 |
| **ICER vs S0 (USD/DALY averted)** | 135.48 | 35.77 | 181.91 | 18.31 | 76.99 | 7.89 | 189.88 | 6.35 | 15.15 | 97.22 | 22.09 | 15.02 | 166.42 |
| **S3. S1 + three-dose catch-up primary at age 18 (HIV+), standard coverage** |  |  |  |  |  |  |  |  |  |  |  |  |  |
| **Discounted total cost (USD)** | 158,596 | 88,913 | 93,466 | 77,768 | 149,085 | 66,352 | 187,008 | 43,317 | 254,845 | 80,268 | 72,811 | 166,522 | 84,182 |
| **Incremental cost vs S0 (USD)** | 1,156 | 997 | 707 | 1,132 | 17,869 | 1,291 | 10,638 | 280 | 2,397 | 376 | 2,384 | 3,977 | 3,090 |
| **ICER vs S0 (USD/DALY averted)** | 283.73 | 141.66 | 296.33 | 22.97 | 246.29 | 25.72 | 393.36 | 21.94 | 30.29 | 77.73 | 55.44 | 39.22 | 528.12 |
| **S4. S2 + three-dose catch-up primary at age 24 (HIV+), standard coverage** |  |  |  |  |  |  |  |  |  |  |  |  |  |
| **Discounted total cost (USD)** | 159,861 | 89,311 | 94,190 | 79,474 | 160,470 | 67,931 | 204,509 | 43,520 | 256,272 | 81,004 | 77,493 | 171,357 | 90,647 |
| **Incremental cost vs S0 (USD)** | 2,422 | 1,394 | 1,431 | 2,838 | 29,253 | 2,869 | 28,140 | 482 | 3,824 | 1,112 | 7,066 | 8,812 | 9,554 |
| **ICER vs S0 (USD/DALY averted)** | 482.27 | 144.24 | 610.42 | 62.75 | 317.24 | 45.14 | 570.13 | 38.60 | 40.03 | 305.34 | 94.63 | 60.02 | 806.71 |
| **S5. S3 with high HIV+ coverage (98%)** |  |  |  |  |  |  |  |  |  |  |  |  |  |
| **Discounted total cost (USD)** | 159,848 | 89,917 | 94,254 | 79,064 | 167,390 | 67,981 | 197,548 | 43,651 | 257,463 | 80,690 | 75,179 | 170,623 | 87,279 |
| **Incremental cost vs S0 (USD)** | 2,408 | 2,000 | 1,495 | 2,428 | 36,173 | 2,919 | 21,178 | 614 | 5,015 | 798 | 4,751 | 8,078 | 6,187 |
| **ICER vs S0 (USD/DALY averted)** | 374.43 | 173.64 | 380.60 | 28.88 | 304.01 | 31.90 | 510.51 | 26.46 | 43.06 | 99.50 | 64.72 | 51.18 | 625.37 |
| **S6. S4 with high HIV+ coverage (98%)** |  |  |  |  |  |  |  |  |  |  |  |  |  |
| **Discounted total cost (USD)** | 162,512 | 90,929 | 95,807 | 82,545 | 191,015 | 71,725 | 232,320 | 44,164 | 260,862 | 82,152 | 84,515 | 180,636 | 100,250 |
| **Incremental cost vs S0 (USD)** | 5,073 | 3,012 | 3,048 | 5,909 | 59,798 | 6,663 | 55,951 | 1,127 | 8,414 | 2,260 | 14,088 | 18,091 | 19,158 |
| **ICER vs S0 (USD/DALY averted)** | 559.38 | 157.27 | 632.45 | 61.17 | 362.03 | 44.08 | 691.38 | 37.21 | 52.78 | 301.12 | 99.87 | 69.05 | 859.29 |

Note: Total costs include discounted vaccine acquisition and delivery costs only; cervical cancer management costs are excluded. Incremental costs and ICERs are calculated relative to S0. ICERs are expressed in USD per discounted DALY averted. Small differences between total and incremental costs may arise from rounding.

**Figure 3.**
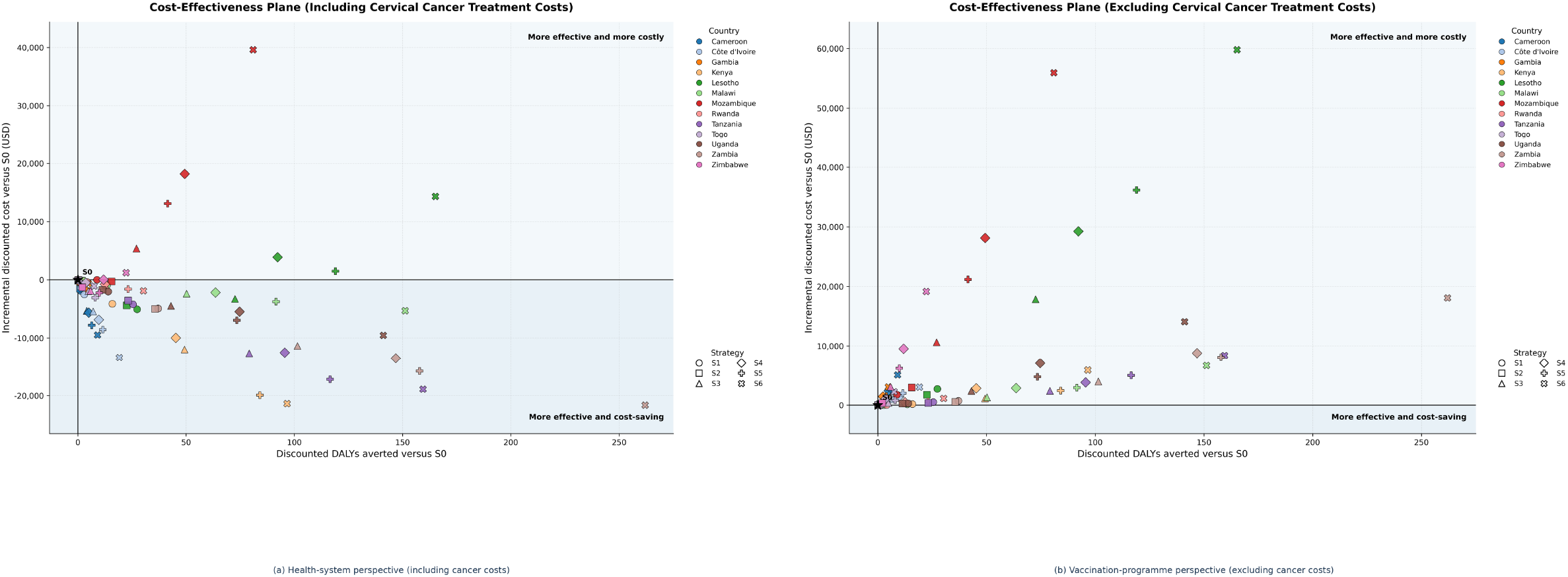
Cost-effectiveness planes of HIV-targeted HPV vaccination strategies across 13 African countries. The horizontal axis represents discounted DALYs averted relative to baseline (S0), while the vertical axis represents incremental discounted costs. Colors identify countries, and marker shapes distinguish vaccination strategies (S1–S6). Points in the lower-right quadrant indicate strategies that are more effective and less costly than S0, whereas points in the upper-right quadrant indicate strategies that are more effective but more costly.

#### 3.3.1 Health-system perspective

When cervical cancer management costs were included, HIV-targeted HPV vaccination was generally more effective and less costly than routine vaccination alone. Of the 78 strategy–country comparisons, 68 were cost-saving relative to S0 (Table 3; Figure 3(a)). In these comparisons, the additional vaccination expenditure was outweighed by reductions in cervical cancer management costs.

Across the 13 countries, the mean total cost under S0 was approximately USD 3.248 million (SD: 1.237 million; range: 1.083– 5.045 million). Mean incremental costs were negative for all six intervention strategies. S5 generated the largest average cost saving, with a mean incremental cost of USD −5,673 (SD: 8,710; range: −19,902 to 13,094), followed by S3 at USD −4,413 (SD: 5,092; range: −12,624 to 5,372). Under S6, the mean incremental cost was USD −3,688 (SD: 16,514; range: −21,621 to 39,609), indicating substantial variation in economic outcomes across countries.

The remaining 10 comparisons were more effective but more costly than S0. In Mozambique, positive incremental costs were observed for S1 and S3–S6, with the largest increase under S6 (USD 39,609). Positive incremental costs were also observed for S4–S6 in Lesotho and for S4 and S6 in Zimbabwe. These comparisons appear in the upper-right quadrant of Figure 3(a), where additional vaccination expenditure exceeded the cervical cancer management costs avoided. Nevertheless, all strategies continued to avert DALYs relative to S0.

Most ICERs were negative under the health-system perspective, indicating that the corresponding strategies were both more effective and less costly than S0. Positive ICERs occurred in the 10 comparisons with additional health-system expenditure (Table 3).

Figure 2(b) further illustrates the variation in net costs per additional vaccine dose. The arithmetic mean marginal net costs were USD −2.65 million for S1 (SD: 2.11 million; range: −8.27 million to 384) and USD −2.14 million for S2 (SD: 1.40 million; range: −4.36 million to −27,320), per 100,000 additional physical vaccine doses. Under S6, the corresponding mean was USD −0.28 million (SD: 0.35 million; range: −1.04 million to 202,864). Booster-only strategies therefore generated larger average savings per additional dose, whereas broader catch-up strategies required more vaccination expenditure and generally produced smaller dose-normalized savings.

#### 3.3.2 Vaccination-programme perspective

When cervical cancer management costs were excluded, all intervention strategies required additional vaccination expenditure relative to S0 (Table 4; Figure 3(b)). The mean incremental programme costs were USD 558 for S1 (SD: 794; range: 34–2,718) and USD 499 for S2 (SD: 872; range: 1–2,953). Combined booster and catch-up strategies required greater expenditure, with S6 generating the highest mean incremental programme cost of USD 15,584 (SD: 19,669; range: 1,127–59,798). These higher costs reflected the additional vaccine doses required for catch-up and booster vaccination.

All intervention strategies remained more effective than S0 and were located in the upper-right quadrant of Figure 3(b). The arithmetic mean direct ICERs were USD 74.16 per DALY averted for S1 (SD: 72.21; range: 8.33–199.20) and USD 74.50 for S2 (SD: 71.35; range: 6.35–189.88). The corresponding mean for S6 was USD 302.08 per DALY averted (SD: 290.70; range: 37.21–859.29). Booster-only strategies therefore had the lowest average direct ICERs, while combined vaccination strategies generally produced larger absolute health gains at greater programme cost.

#### 3.3.3 Sequential cost-effectiveness analysis

Under the health-system perspective, positive sequential ICERs remained in only four countries after excluding dominated strategies (Table A6). In Lesotho, sequential ICERs ranged from USD 39.85 to USD 278.05 per additional DALY averted, while in Mozambique they ranged from USD 491.96 to USD 678.70. The sequential ICER for S6 relative to S5 was USD 1,033.69 in Gambia and USD 279.69 in Zimbabwe. In the remaining nine countries, only one strategy remained on the cost-effectiveness frontier, and no sequential ICER was applicable.

Under the vaccination-programme perspective, multiple non-dominated strategies remained in all 13 countries (Table A7). Sequential ICERs ranged from USD 6.35 per additional DALY averted for S2 relative to S0 in Rwanda to USD 1,742.90 for S6 relative to S5 in Gambia. Lower sequential ICERs were generally observed in countries such as Rwanda, Malawi, and Kenya, whereas higher values occurred when additional vaccination expenditure produced relatively small incremental health gains. In Togo, S5 was the most effective non-dominated strategy, while S6 was strictly dominated because it generated fewer DALYs averted at a higher cost.

### 3.4 Sensitivity analysis

We assessed the robustness of model outcomes to uncertainty in calibrated epidemiological parameters using a deterministic one-way sensitivity analysis for Mozambique. Each parameter was varied independently by ±20% around its calibrated value, and changes in total discounted DALYs, health-system costs, and vaccine doses under S0 were summarized using tornado diagrams (Figure 4).

**Figure 4.**
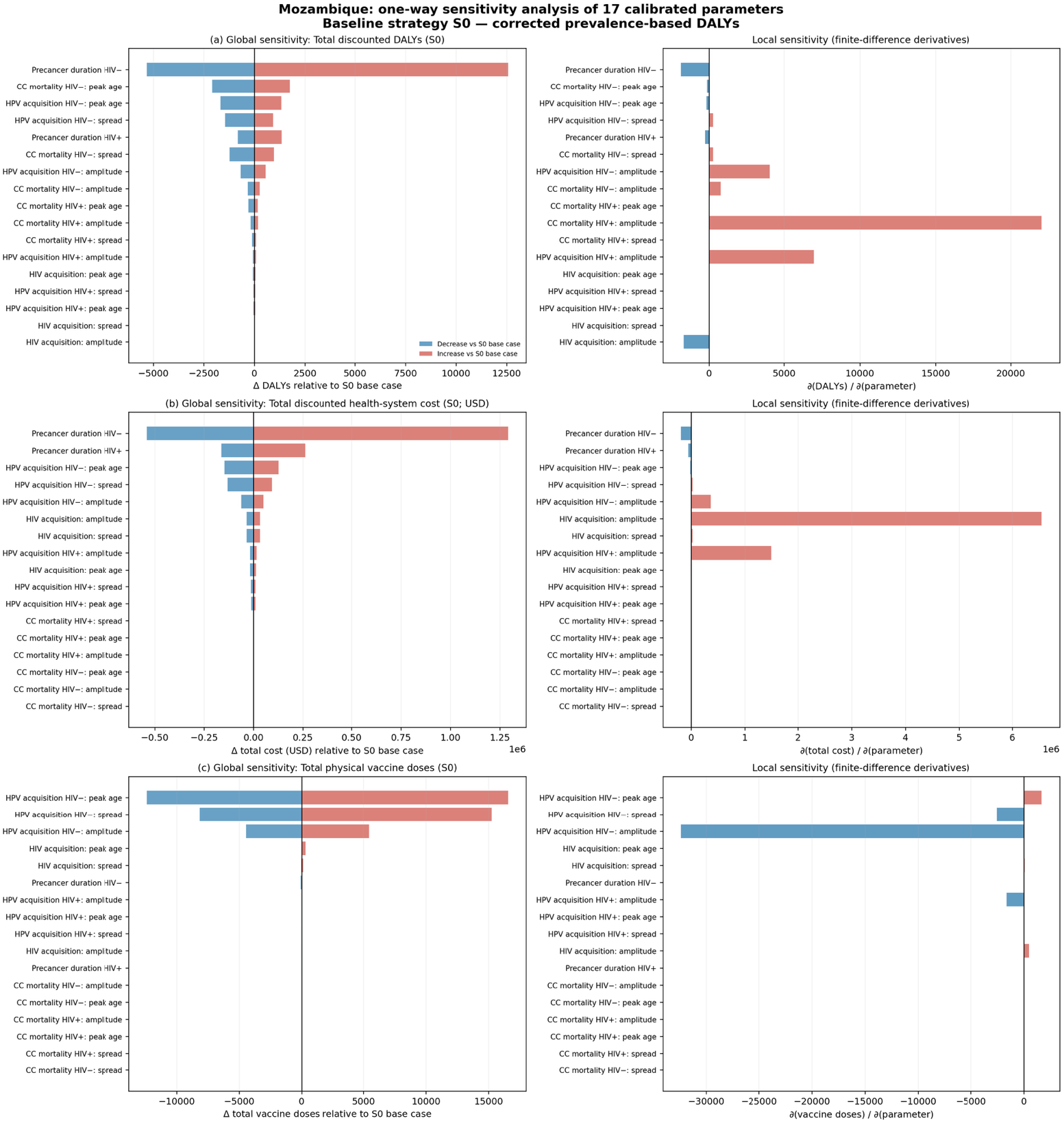
One-way sensitivity analysis of the 17 calibrated epidemiological parameters in Mozambique under baseline vaccination (S0). Left panels show minimum and maximum changes in (a) total discounted DALYs, (b) total discounted health-system costs and (c) physical vaccine doses. Right panels show local finite-difference sensitivity derivatives.

Sensitivity analysis varying cervical cancer treatment costs between 50% and 150% of the base-case value showed that higher treatment costs reduced incremental health-system expenditure, although the combined vaccination strategies S3–S6 remained more costly than S0 across the evaluated range (Figure A4).

The largest variation in discounted DALYs and total costs was driven by the duration of precancerous disease among HIV-negative women (Figure 4a–b). Total vaccine doses were primarily influenced by parameters governing the age pattern of HPV acquisition among HIV-negative women, particularly peak age and spread (Figure 4c).

The probabilistic sensitivity analysis showed that all vaccination strategies remained health-improving relative to S0 across 2,000 simulations (Figure A3). Under ±10% variation in calibrated epidemiological parameters, S1 and S2 were cost-saving in 50.0% and 63.8% of simulations, respectively, whereas S3–S6 consistently incurred additional health-system costs. These findings indicate that the health benefits were robust to the parameter uncertainty examined, although cost-saving conclusions varied by strategy.

## 4. Discussion

This study evaluated the health impact and cost-effectiveness of HIV-targeted HPV vaccination strategies across 13 African countries. All strategies provided additional health benefits beyond routine vaccination, although their relative advantages depended on the programme objective. Combined booster and catch-up vaccination, particularly the high-coverage strategy at age 24 years (S6), generated the greatest average absolute health gains, while booster-only strategies (S1 and S2) achieved the highest health gains per additional vaccine dose. Most strategies were cost-saving when avoided cervical cancer management costs were included, but required additional vaccination-programme expenditure. These findings highlight the importance of balancing total health impact, dose efficiency, and affordability when selecting vaccination strategies.

The larger health gains achieved by combined strategies reflect their ability to reach previously unvaccinated women living with HIV in addition to providing boosters to those vaccinated during adolescence. However, these strategies also require more vaccine doses, explaining their lower dose-normalized efficiency compared with booster-only approaches. This distinction is particularly relevant where vaccine supply is limited, as maximizing population health benefits may require a different strategy from maximizing benefits per dose [13]. Vaccination at age 24 generally produced greater health gains among the combined strategies, but this pattern was not consistent for booster-only vaccination. Differences between target ages reflect the number of eligible women, previous HPV exposure, remaining susceptibility, and duration of protection rather than greater effectiveness when vaccination is delivered closer to cervical cancer onset. HPV vaccines prevent new infections but do not treat established disease [5, 4].

The magnitude of health gains varied substantially across countries despite common vaccination coverage assumptions. This variation reflects differences in HIV epidemiology, cervical cancer burden, and the age distribution of women eligible for vaccination. Women living with HIV experience a disproportionately high risk of HPV-related disease, although the burden of cervical cancer associated with HIV varies considerably between settings [1, 2]. Country-specific epidemiological characteristics are therefore important when estimating the potential benefits of HIV-targeted vaccination.

Economic outcomes further emphasized the importance of the analytic perspective. When cervical cancer management costs were included, most strategies generated health gains alongside cost savings, although additional costs remained in some settings, particularly Mozambique. When these costs were excluded, all strategies required additional vaccination expenditure. This difference has practical implications because immunization programmes bear the immediate costs of vaccine procurement and delivery, while savings from prevented cervical cancer arise later and may benefit other health services. Consequently, lifetime cost-effectiveness does not necessarily imply short-term affordability, and complementary budget impact analyses would be needed to inform implementation decisions [14]. Actual delivery costs may also vary with programme scale, staffing, and delivery platforms [10].

The proposed booster strategies should be interpreted as exploratory interventions rather than established vaccination recommendations. WHO recommends at least two HPV vaccine doses for individuals living with HIV, with three doses where feasible, but does not currently recommend routine booster vaccination following completion of primary vaccination [4]. Although available studies support the safety and immunogenicity of HPV vaccines among people living with HIV, evidence regarding long-term clinical protection remains limited [6, 15]. The PAPILLON trial demonstrated non-inferior short-term seroconversion with two compared with three doses among women with well-controlled HIV, but did not establish long-term clinical effectiveness or the benefits of additional booster vaccination [16]. Further evidence is therefore needed to support the booster assumptions evaluated in this model.

From an implementation perspective, HIV-care services could provide opportunities to deliver targeted HPV vaccination during diagnosis, treatment initiation, or routine follow-up. However, identifying previous HPV vaccination status may be difficult where immunization records are incomplete or poorly linked to HIV services. Experience integrating cervical cancer prevention into HIV care suggests that service integration may be feasible, although staffing, supply chains, and continuity of care remain important challenges [17]. Moreover, the high coverage assumed under S5 and S6 should be considered an aspirational scenario rather than an estimate of achievable uptake.

Equity is another important consideration. Targeting women living with HIV directs additional preventive resources towards a population with increased cervical cancer risk. Nevertheless, programmes delivered exclusively through HIV clinics may fail to reach women with undiagnosed HIV, limited access to healthcare, or barriers related to geography and stigma. In the model, vaccination eligibility was based on HIV status without explicitly accounting for diagnosis or engagement with care. Effective implementation would therefore require approaches that improve accessibility among underserved populations, while maintaining routine HPV vaccination for all eligible adolescents [18].

Several limitations should be acknowledged. The closed-cohort model did not capture HPV transmission or indirect herd-protection effects, while screening, cancer recurrence, and detailed HIV-related clinical heterogeneity were not explicitly represented. Uniform vaccination coverage assumptions improved comparability across countries but did not reflect differences in national programme uptake. Uncertainty also remains regarding vaccine efficacy, duration of protection, and booster-induced immunity among women living with HIV. The probabilistic sensitivity analysis, conducted for Mozambique, examined uncertainty in calibrated epidemiological parameters but did not vary all vaccination and economic inputs. Furthermore, country-specific cervical cancer management costs were partly estimated from economic proxies, which may not fully capture differences in treatment availability and healthcare delivery.

Overall, HIV-targeted HPV vaccination could complement routine adolescent immunization and reduce cervical cancer burden among women living with HIV. The preferred approach will depend on local epidemiology, vaccine availability, programme affordability, and the capacity to reach eligible women. Further evidence on booster effectiveness, duration of protection, and feasible delivery through HIV services is needed to inform future vaccination policy.

## Data Availability

All epidemiological and demographic input data used in this study were obtained from publicly available sources, including the Global Burden of Disease Study 2023, as cited in the manuscript.

## Author Contributions

SBM and ST contributed to the study conception and design, developed the initial modelling framework, and prepared the first draft of the manuscript. SBM subsequently revised and extended the modelling code, performed the final simulations and analyses, and substantially rewrote the manuscript. KA and TAH critically reviewed the methodology, results, and manuscript drafts and provided important intellectual input. SBM, AK and CT supervised the study and provided overall scientific guidance throughout the project. All authors reviewed and approved the final version of the manuscript.

