## Supplementary material for "Optimizing HPV Vaccination for Cervical Cancer Prevention in Women Living with HIV: A Cost-Effectiveness Analysis Across 13 African Countries": supplemental File.pdf

### Appendices

#### Appendix 1. Model structure and mathematical formulation

We developed an age-structured, annual-cycle state-transition model to simulate the natural history of oncogenic HPV infection and cervical cancer in a closed female cohort stratified by HIV status (Figure 1). A cohort of 100,000 females was followed from age 0 to 100 years, or until death, without births or migration during follow-up.

Parallel health states represented HIV-negative and HIV-positive individuals, accounting for differences in HPV acquisition, clearance, disease progression, and mortality. The two strata were linked through age-dependent HIV acquisition, allowing transitions from HIV-negative to corresponding HIV-positive health states.

### 1.1 Health states and state variables

For HIV-negative individuals, the model includes susceptible unvaccinated ($S$), vaccinated ($V$), HPV-infected ($I$), three sequential precancer states ($P_1$, $P_2$, and $P_3$), invasive cervical cancer ($C$), and treated cancer survivors ($R$). The three precancer states were used as tunnel compartments to represent the time spent in precancer before progression to invasive cervical cancer. Individuals in any precancer state could either progress to the next stage or clear the lesion and return to the susceptible state.

The HIV-positive structure contains the corresponding states ($S^+$, $V^+$, $I^+$, $P_1^+$, $P_2^+$, $P_3^+$, $C^+$, and $R^+$), together with a boosted-vaccination state ($V^{b+}$). This additional state represents previously vaccinated women living with HIV who receive a booster dose.

Cervical cancer survivors enter $R$ or $R^+$ following recovery from invasive cancer. These states do not return to the HPV natural-history pathway, although individuals remain subject to background mortality and, for HIV-positive survivors, HIV-related mortality. HIV-negative survivors may also acquire HIV and move from $R$ to $R^+$.

Deaths are recorded in separate cumulative compartments according to cause and HIV status. These comprise cervical cancer deaths among HIV-negative ($D_c$) and HIV-positive ($D_c^+$) individuals, background deaths among HIV-negative ($D$) and HIV-positive ($D^+$) individuals, and HIV-related deaths among HIV-positive individuals ($D_H^+$).

### 1.2 Core parameters

To represent differences in HPV natural history by HIV status, separate parameter sets were used for HIV-negative and HIV-positive individuals. These HIV-specific parameters allowed HPV acquisition, clearance, precancer progression, cervical cancer recovery, and cervical cancer mortality to differ by HIV status, with lower HPV clearance and poorer cervical cancer outcomes assumed among women living with HIV.

HPV acquisition among susceptible individuals is governed by the age-specific hazards $\lambda_{\mathrm{HPV}}(a)$ for HIV-negative women and $\lambda_{\mathrm{HPV}}^+(a)$ for women living with HIV. Following infection, HPV may clear at rates $c_{\mathrm{HPV}}$ and $c_{\mathrm{HPV}}^+$, respectively, with individuals returning to the corresponding susceptible state:

$$I \to S \qquad \text{and} \qquad I^+ \to S^+.$$

No durable natural immunity was assumed following clearance, so individuals could subsequently be reinfected.

Progression from HPV infection to the first precancer stage is governed by $\lambda_{IP_1}$ for HIV-negative individuals and $\lambda_{IP_1}^+$ for HIV-positive individuals. Precancer progression then occurs sequentially through the transitions

$$P_1 \to P_2, \qquad P_2 \to P_3, \qquad P_3 \to C,$$

with corresponding HIV-specific transition rates. Each precancer state may also clear directly to susceptibility, with rates $c_{P_1}$, $c_{P_2}$, and $c_{P_3}$ for HIV-negative individuals and $c_{P_1}^+$, $c_{P_2}^+$, and $c_{P_3}^+$ for HIV-positive individuals.

Individuals with invasive cervical cancer may recover and enter the post-treatment survivor state at rates $r_c$ and $r_c^+$:

$$C \to R \qquad \text{and} \qquad C^+ \to R^+.$$

Cancer recurrence and subsequent post-treatment disease progression were not modelled explicitly.

HIV acquisition is governed by the age-specific annual hazard $\lambda_{\mathrm{HIV}}(a)$ and may occur from any living HIV-negative health state. HIV acquisition does not reset HPV or cervical cancer history. Instead, individuals move to the corresponding HIV-positive state while retaining their current disease stage, for example

$$S \to S^+, \quad V \to V^+, \quad I \to I^+, \quad P_j \to P_j^+, \quad C \to C^+,$$

where $j = 1,2,3$. After this transition, HIV-specific HPV progression, clearance, cancer recovery, and mortality rates apply.

Mortality includes age-specific background mortality $\mu(a)$ for all living individuals, additional HIV-related mortality $\mu_H(a)$ among HIV-positive individuals, and cervical cancer-specific mortality $\mu_C(a)$ and $\mu_C^+(a)$ for HIV-negative and HIV-positive cancer cases, respectively. Population conservation was checked at each annual cycle across all living and death states.

Vaccination is represented through the vaccinated susceptible states $V$ and $V^+$ and the boosted state $V^{b+}$ among women living with HIV. Vaccination is prophylactic and reduces only the hazard of HPV acquisition; it does not alter clearance or progression after infection has occurred. Primary vaccination uptake is represented by the age- and time-dependent functions $\gamma(a,t)$ for HIV-negative individuals and $\gamma_h(a,t)$ for HIV-positive individuals, while booster uptake is represented by $\rho(a,t)$. Vaccine protection is implemented through the residual HPV acquisition risks $\delta(a,t)$, $\delta_h(a,t)$, and $\delta_b(a,t)$ for HIV-negative, HIV-positive, and boosted HIV-positive individuals, respectively. Booster vaccination is represented by the transition

$$V^+ \to V^{b+},$$

with vaccine protection restored and the duration of protection reset according to the booster assumptions (Table A1).

Table A1. Model parameters and standardized notation.

| Parameter | Description |
|---|---|
| $\lambda_{\mathrm{HPV}}(a)$ | Age-specific HPV acquisition hazard among HIV-negative individuals. |
| $\lambda_{\mathrm{HPV}}^+(a)$ | Age-specific HPV acquisition hazard among HIV-positive individuals. |
| $\lambda_{\mathrm{HIV}}(a)$ | Age-specific HIV acquisition hazard. |
| $c_{\mathrm{HPV}}$ | HPV clearance rate among HIV-negative individuals ($I \to S$). |
| $c_{\mathrm{HPV}}^+$ | HPV clearance rate among HIV-positive individuals ($I^+ \to S^+$). |
| $\lambda_{IP_1}$ | Progression rate from infection to the first precancer state ($I \to P_1$). |
| $\lambda_{IP_1}^+$ | Progression rate from infection to the first precancer state among HIV-positive individuals ($I^+ \to P_1^+$). |
| $\lambda_{P_1P_2}, \lambda_{P_2P_3}, \lambda_{P_3C}$ | HIV-negative progression rates through the precancer tunnel states and to invasive cancer. |
| $\lambda_{P_1P_2}^+, \lambda_{P_2P_3}^+, \lambda_{P_3C}^+$ | HIV-positive progression rates through the precancer tunnel states and to invasive cancer. |
| $c_{P_1}, c_{P_2}, c_{P_3}$ | Clearance rates from precancer states to susceptibility among HIV-negative individuals. |
| $c_{P_1}^+, c_{P_2}^+, c_{P_3}^+$ | Clearance rates from precancer states to susceptibility among HIV-positive individuals. |
| $r_C$ | Recovery rate from invasive cancer to the survivor state ($C \to R$). |
| $r_C^+$ | Recovery rate from invasive cancer among HIV-positive individuals ($C^+ \to R^+$). |
| $\mu(a)$ | Age-specific background mortality rate. |
| $\mu_H(a)$ | Age-specific HIV-related mortality rate. |
| $\mu_C(a)$ | Cervical cancer mortality rate among HIV-negative individuals. |
| $\mu_C^+(a)$ | Cervical cancer mortality rate among HIV-positive individuals. |
| $\gamma(a,t)$ | Primary vaccination uptake among HIV-negative individuals. |
| $\gamma_h(a,t)$ | Primary or catch-up vaccination uptake among HIV-positive individuals. |
| $\rho(a,t)$ | Booster uptake among previously vaccinated HIV-positive individuals. |
| $VE(\tau)$ | Vaccine efficacy among HIV-negative individuals at time $\tau$ since vaccination. |
| $VE^+(\tau)$ | Vaccine efficacy among HIV-positive individuals at time $\tau$ since primary vaccination. |
| $VE^{b+}(\tau)$ | Vaccine efficacy among HIV-positive individuals at time $\tau$ since booster vaccination. |
| $\delta(a,t), \delta_h(a,t), \delta_b(a,t)$ | Residual HPV acquisition risks after primary or booster vaccination. |

The fixed natural-history parameters used in all country models are shown in Table A2. Parameter values were specified as annual probabilities and converted to continuous-time rates using

$$r = -\ln(1-p),$$

before being applied in the competing-risks transition equations.

Table A2. Fixed HPV and cervical cancer natural-history parameters used in the final model.

| Parameter | Description | Annual probability | Annual rate | Source / basis |
|---|---|---|---|---|
| $\lambda_{IP_1}$ | Progression from HPV infection to the first precancer state among HIV-negative individuals | 0.040 | 0.0408 | Assumed; informed by published HPV natural-history modelling estimates [19] |
| $\lambda_{IP_1}^{+}$ | Progression from HPV infection to the first precancer state among HIV-positive individuals | 0.080 | 0.0834 | Assumed; informed by published HPV–HIV modelling and cohort evidence [19, 20] |
| $c_{\text{HPV}}$ | HPV clearance among HIV-negative individuals ($I \to S$) | 0.300 | 0.3567 | Selected within published HPV-clearance estimates [19, 21] |
| $c_{\text{HPV}}^{+}$ | HPV clearance among HIV-positive individuals ($I^{+} \to S^{+}$) | 0.212 | 0.2383 | Published model estimate [19]; supported by HIV-positive natural-history evidence [20] |
| $c_{P_1}$ | Clearance from the first precancer tunnel state among HIV-negative individuals ($P_1 \to S$) | 0.100 | 0.1054 | Assumed; informed by evidence on regression of cervical precancerous lesions [22, 23] |
| $c_{P_2}$ | Clearance from the second precancer tunnel state among HIV-negative individuals ($P_2 \to S$) | 0.080 | 0.0834 | Assumed; informed by cervical lesion-regression evidence [22, 23] |
| $c_{P_3}$ | Clearance from the third precancer tunnel state among HIV-negative individuals ($P_3 \to S$) | 0.050 | 0.0513 | Assumed; lower clearance was assigned to the later precancer tunnel state, informed by lesion natural-history evidence [22, 23] |
| $c_{P_1}^{+}$ | Clearance from the first precancer tunnel state among HIV-positive individuals ($P_1^{+} \to S^{+}$) | 0.070 | 0.0726 | Assumed; lower than the corresponding HIV-negative clearance probability to reflect greater lesion persistence among WLHIV [20, 24] |
| $c_{P_2}^{+}$ | Clearance from the second precancer tunnel state among HIV-positive individuals ($P_2^{+} \to S^{+}$) | 0.060 | 0.0619 | Assumed; informed by evidence of reduced regression and greater progression among WLHIV [20, 24] |
| $c_{P_3}^{+}$ | Clearance from the third precancer tunnel state among HIV-positive individuals ($P_3^{+} \to S^{+}$) | 0.040 | 0.0408 | Assumed; lower clearance in the later precancer state reflects greater persistence of cervical disease among WLHIV [20, 24] |
| $r_C$ | Recovery from invasive cervical cancer among HIV-negative individuals ($C \to R$) | 0.151 | 0.1637 | Published model estimate [19] |
| $r_C^{+}$ | Recovery from invasive cervical cancer among HIV-positive individuals ($C^{+} \to R^{+}$) | 0.068 | 0.0704 | Assumed; a lower recovery probability was assigned to HIV-positive individuals, informed by evidence of poorer cancer survival among people living with HIV [25]. The numerical value was specified as a modelling assumption. |

Notes: Annual probabilities were converted to continuous-time rates using $r = -\ln(1 - p)$. The three precancer states ($P_1$–$P_3$) are sequential tunnel states used to represent time spent in precancer and should not be interpreted as direct representations of CIN1, CIN2, and CIN3. Exact empirical estimates corresponding to the model-specific tunnel-state clearance probabilities were not available. These parameters were therefore specified as fixed modelling assumptions informed by published evidence on cervical lesion regression and the greater persistence and progression of cervical disease among women living with HIV. Progression rates between successive precancer tunnel states were not fixed in this table; they were derived from the calibrated total precancer duration separately for HIV-negative and HIV-positive individuals.

### 1.3 Model equations

The model is a static, age-structured cohort state-transition model with annual cycles. HPV acquisition hazards are specified independently of the number of infected individuals in the cohort. Consequently, HPV transmission and indirect protection through herd immunity are not represented.

#### 1.3.1 States and registers

The living states for HIV-negative individuals are

$$S,\ V,\ I,\ P_1,\ P_2,\ P_3,\ C,\ R,$$

where $S$ and $V$ denote susceptible unvaccinated and vaccinated individuals, $I$ denotes HPV infection, $P_1$–$P_3$ are sequential precancer tunnel states, $C$ denotes invasive cervical cancer, and $R$ denotes post-treatment cancer survival. Cumulative cervical cancer and background deaths are recorded in $D_c$ and $D$, respectively.

The corresponding living states for HIV-positive individuals are

$$S^{+},\ V^{+},\ V^{b+},\ I^{+},\ P_1^{+},\ P_2^{+},\ P_3^{+},\ C^{+},\ R^{+},$$

where $V^{b+}$ denotes the boosted vaccinated state. Cumulative cervical cancer, background, and HIV-related deaths are recorded in $D_c^{+}$, $D^{+}$, and $D_H^{+}$, respectively.

#### 1.3.2 Competing-risk transition probabilities

Transitions are implemented using a competing-risks formulation. Let $h_{xk}(a,t)$ denote the hazard of moving from state $x$ to destination $k$, and let

$$H_x(a,t) = \sum_k h_{xk}\,(a,t)$$

be the total outgoing hazard from state $x$. For an annual cycle of length $\Delta t = 1$, the probability of remaining in state $x$ is

$$q_x(a,t) = \exp\{-H_x(a,t)\},$$

and the probability of moving from $x$ to destination $k$ is

$$p_{xk}(a,t) = \frac{h_{xk}(a,t)}{H_x(a,t)}[1 - \exp\{-H_x(a,t)\}].$$

If $H_x(a,t) = 0$, then $q_x(a,t) = 1$ and all outgoing transition probabilities are zero. This formulation ensures that competing events are combined consistently and that the total probability of remaining in or leaving a state equals one.

#### 1.3.3 HIV-negative transition hazards

For HIV-negative individuals, the total outgoing hazards are

$$\begin{aligned}
H_S &= \gamma(a,t) + \lambda_{\mathrm{HPV}}(a) + \lambda_{\mathrm{HIV}}(a) + \mu(a),\\
H_V &= \delta(a,t)\lambda_{\mathrm{HPV}}(a) + \lambda_{\mathrm{HIV}}(a) + \mu(a),\\
H_I &= c_{\mathrm{HPV}} + \lambda_{IP_1} + \lambda_{\mathrm{HIV}}(a) + \mu(a),\\
H_{P_1} &= c_{P_1} + \lambda_{P_1P_2} + \lambda_{\mathrm{HIV}}(a) + \mu(a),\\
H_{P_2} &= c_{P_2} + \lambda_{P_2P_3} + \lambda_{\mathrm{HIV}}(a) + \mu(a),\\
H_{P_3} &= c_{P_3} + \lambda_{P_3C} + \lambda_{\mathrm{HIV}}(a) + \mu(a),\\
H_C &= r_C + \mu_C(a) + \lambda_{\mathrm{HIV}}(a) + \mu(a),\\
H_R &= \lambda_{\mathrm{HIV}}(a) + \mu(a).
\end{aligned}$$

Here, $\gamma(a,t)$ is the primary vaccination hazard and $\delta(a,t)$ is the residual HPV acquisition risk among vaccinated HIV-negative individuals.

Using the competing-risk probabilities defined above, the HIV-negative states are updated as

$$\begin{aligned}
S_{t+1} &= q_S S_t + p_{I\to S} I_t + p_{P_1\to S} P_{1,t} + p_{P_2\to S} P_{2,t} + p_{P_3\to S} P_{3,t},\\
V_{t+1} &= q_V V_t + p_{S\to V} S_t,\\
I_{t+1} &= q_I I_t + p_{S\to I} S_t + p_{V\to I} V_t,\\
P_{1,t+1} &= q_{P_1} P_{1,t} + p_{I\to P_1} I_t,\\
P_{2,t+1} &= q_{P_2} P_{2,t} + p_{P_1\to P_2} P_{1,t},\\
P_{3,t+1} &= q_{P_3} P_{3,t} + p_{P_2\to P_3} P_{2,t},\\
C_{t+1} &= q_C C_t + p_{P_3\to C} P_{3,t},\\
R_{t+1} &= q_R R_t + p_{C\to R} C_t.
\end{aligned}$$

New cervical cancer cases and primary vaccinations are recorded as

$$\begin{aligned}
\text{new_cases}_t &= p_{P_3\to C} P_{3,t},\\
\text{new_vax}_t &= p_{S\to V} S_t.
\end{aligned}$$

The cumulative HIV-negative death registers are updated as

$$\begin{aligned}
D_{c,t+1} &= D_{c,t} + p_{C\to D_c} C_t,\\
D_{t+1} &= D_t + p_{S\to D} S_t + p_{V\to D} V_t + p_{I\to D} I_t + p_{P_1\to D} P_{1,t} + p_{P_2\to D} P_{2,t} + p_{P_3\to D} P_{3,t} + p_{C\to D} C_t + p_{R\to D} R_t.
\end{aligned}$$

#### 1.3.4 HIV-positive transition hazards

For HIV-positive individuals, the total outgoing hazards are

$$
\begin{aligned}
H_{S^+} &= \gamma_h(a,t) + \lambda^+_{\mathrm{HPV}}(a) + \mu_H(a) + \mu(a),\\
H_{V^+} &= \rho(a,t) + \delta_h(a,t)\lambda^+_{\mathrm{HPV}}(a) + \mu_H(a) + \mu(a),\\
H_{V^{b+}} &= \delta_b(a,t)\lambda^+_{\mathrm{HPV}}(a) + \mu_H(a) + \mu(a),\\
H_{I^+} &= c^+_{\mathrm{HPV}} + \lambda^+_{IP_1} + \mu_H(a) + \mu(a),\\
H_{P_1^+} &= c^+_{P_1} + \lambda^+_{P_1P_2} + \mu_H(a) + \mu(a),\\
H_{P_2^+} &= c^+_{P_2} + \lambda^+_{P_2P_3} + \mu_H(a) + \mu(a),\\
H_{P_3^+} &= c^+_{P_3} + \lambda^+_{P_3C} + \mu_H(a) + \mu(a),\\
H_{C^+} &= r^+_C + \mu^+_C(a) + \mu_H(a) + \mu(a),\\
H_{R^+} &= \mu_H(a) + \mu(a).
\end{aligned}
$$

Here, $\gamma_h(a,t)$ is the primary or catch-up vaccination hazard among HIV-positive susceptible individuals, $\rho(a,t)$ is the booster hazard, and $\delta_h(a,t)$ and $\delta_b(a,t)$ are the residual HPV acquisition risks among vaccinated and boosted HIV-positive individuals, respectively.

The HIV-positive states receive both within-stratum flows and HIV acquisition flows from corresponding HIV-negative states:

$$
\begin{aligned}
S^+_{t+1} &= q_{S^+}S^+_t + p_{S\to S^+}S_t + p_{I^+\to S^+}I^+_t + p_{P_1^+\to S^+}P^+_{1,t} + p_{P_2^+\to S^+}P^+_{2,t} + p_{P_3^+\to S^+}P^+_{3,t},\\
V^+_{t+1} &= q_{V^+}V^+_t + p_{S^+\to V^+}S^+_t + p_{V\to V^+}V_t,\\
V^{b+}_{t+1} &= q_{V^{b+}}V^{b+}_t + p_{V^+\to V^{b+}}V^+_t,\\
I^+_{t+1} &= q_{I^+}I^+_t + p_{S^+\to I^+}S^+_t + p_{V^+\to I^+}V^+_t + p_{V^{b+}\to I^+}V^{b+}_t + p_{I\to I^+}I_t,\\
P^+_{1,t+1} &= q_{P_1^+}P^+_{1,t} + p_{I^+\to P_1^+}I^+_t + p_{P_1\to P_1^+}P_{1,t},\\
P^+_{2,t+1} &= q_{P_2^+}P^+_{2,t} + p_{P_1^+\to P_2^+}P^+_{1,t} + p_{P_2\to P_2^+}P_{2,t},\\
P^+_{3,t+1} &= q_{P_3^+}P^+_{3,t} + p_{P_2^+\to P_3^+}P^+_{2,t} + p_{P_3\to P_3^+}P_{3,t},\\
C^+_{t+1} &= q_{C^+}C^+_t + p_{P_3^+\to C^+}P^+_{3,t} + p_{C\to C^+}C_t,\\
R^+_{t+1} &= q_{R^+}R^+_t + p_{C^+\to R^+}C^+_t + p_{R\to R^+}R_t.
\end{aligned}
$$

New cervical cancer cases, primary vaccinations, and booster vaccinations among HIV-positive individuals are recorded as

$$
\begin{aligned}
\text{new\_cases}^+_t &= p_{P_3^+\to C^+}P^+_{3,t},\\
\text{new\_vax}^+_t &= p_{S^+\to V^+}S^+_t,\\
\text{new\_boost}^+_t &= p_{V^+\to V^{b+}}V^+_t.
\end{aligned}
$$

The HIV-positive cumulative death registers are updated as

$$
\begin{aligned}
D^+_{c,t+1} &= D^+_{c,t} + p_{C^+\to D_c^+}C^+_t,\\
D^+_{t+1} &= D^+_t + p_{S^+\to D^+}S^+_t + p_{V^+\to D^+}V^+_t + p_{V^{b+}\to D^+}V^{b+}_t\\
&\quad + p_{I^+\to D^+}I^+_t + p_{P_1^+\to D^+}P^+_{1,t} + p_{P_2^+\to D^+}P^+_{2,t}\\
&\quad + p_{P_3^+\to D^+}P^+_{3,t} + p_{C^+\to D^+}C^+_t + p_{R^+\to D^+}R^+_t,\\
D^+_{H,t+1} &= D^+_{H,t} + p_{S^+\to D_H^+}S^+_t + p_{V^+\to D_H^+}V^+_t + p_{V^{b+}\to D_H^+}V^{b+}_t\\
&\quad + p_{I^+\to D_H^+}I^+_t + p_{P_1^+\to D_H^+}P^+_{1,t} + p_{P_2^+\to D_H^+}P^+_{2,t}\\
&\quad + p_{P_3^+\to D_H^+}P^+_{3,t} + p_{C^+\to D_H^+}C^+_t + p_{R^+\to D_H^+}R^+_t.
\end{aligned}
$$

#### 1.3.5 Population conservation

Let $N_t$ denote the sum of all living states and cumulative death registers in both HIV strata. The competing-risks formulation conserves the cohort population at each annual cycle, up to numerical precision:

$$N_{t+1} = N_t.$$

The variables new_cases, new_vax, and new_boost are flow counters and are not included as population stocks in the conservation equation.

### Appendix 2. Calibration objective and optimization

The model was calibrated separately for each country by comparing simulated outputs with three age-specific epidemiological targets: cervical cancer incidence, cervical cancer mortality, and HIV prevalence. At each evaluation of the objective function, the model was simulated over the full age range, and its annual outputs were aggregated to the same age classes as the observed data. Cervical cancer incidence and mortality were calculated across the combined HIV-negative and HIV-positive populations because the observed country-level estimates were not separated by HIV status.

#### 2.1 Calibration parameters

For each country, calibration estimated a 17-dimensional parameter vector,

$$\boldsymbol{\theta} = \left(\boldsymbol{\theta}_{\mathrm{HPV}}, \boldsymbol{\theta}_{\mathrm{HPV}}^{+}, \boldsymbol{\theta}_{\mu_C}, \boldsymbol{\theta}_{\mu_C}^{+}, \boldsymbol{\theta}_{\mathrm{HIV}}, T_P, T_P^{+}\right).$$

Five age-dependent hazards were represented by Gaussian functions: the HPV acquisition hazards for HIV-negative and HIV-positive individuals, the cervical cancer mortality hazards for HIV-negative and HIV-positive individuals, and the HIV acquisition hazard. For a generic age-dependent hazard $f(a)$, the Gaussian form was

$$f(a; A, m, s) = A\exp\left[-\frac{(a-m)^2}{2s^2}\right],$$

where $A$ is the amplitude, $m$ is the age at which the function reaches its maximum, and $s$ controls its dispersion. Each function therefore contributed three estimated parameters, giving 15 Gaussian parameters in total.

The remaining two calibrated parameters, $T_P$ and $T_P^{+}$, represent the total duration of the three-stage precancer pathway among HIV-negative and HIV-positive individuals, respectively. Equal expected durations were assumed for the three tunnel states within each HIV stratum. The transition rate from each precancer stage was derived from the corresponding total duration and the fixed clearance rate. For example,

$$\lambda_{P_j P_{j+1}} = \frac{3}{T_P} - c_{P_j}, \qquad j = 1,2,$$

and

$$\lambda_{P_3 C} = \frac{3}{T_P} - c_{P_3}.$$

The equivalent HIV-positive progression rates were calculated using $T_P^{+}$ and the HIV-positive precancer clearance rates.

#### 2.2 Normalized model–data errors

Let $y_{kg}$ denote the observed value for outcome $k$ in age group $g$, and let $\hat{y}_{kg}(\boldsymbol{\theta})$ denote the corresponding model prediction. The three calibration outcomes were $k \in \{\mathrm{inc}, \mathrm{mort}, \mathrm{hiv}\}$, representing cervical cancer incidence, cervical cancer mortality, and HIV prevalence.

For each outcome, the discrepancy between observed and simulated values was calculated using a mean-normalized mean squared error:

$$E_k(\boldsymbol{\theta}) = \frac{1}{n_k}\sum_{g=1}^{n_k}\left[\frac{\hat{y}_{kg}(\boldsymbol{\theta}) - y_{kg}}{\max\left(n_k^{-1}\sum_{g=1}^{n_k}|y_{kg}|,\ 10^{-9}\right)}\right]^2.$$

Normalization by the mean absolute observed value placed the three outcomes on comparable scales. The weighted model–data error was

$$E_{\mathrm{data}}(\boldsymbol{\theta}) = w_{\mathrm{inc}}E_{\mathrm{inc}}(\boldsymbol{\theta}) + w_{\mathrm{mort}}E_{\mathrm{mort}}(\boldsymbol{\theta}) + w_{\mathrm{hiv}}E_{\mathrm{hiv}}(\boldsymbol{\theta}),$$

with

$$w_{\mathrm{inc}} = 1, \qquad w_{\mathrm{mort}} = 1, \qquad w_{\mathrm{hiv}} = 5.$$

The larger weight assigned to HIV prevalence was used to improve agreement with the observed age-specific HIV distribution.

#### 2.3 Regularization and biological plausibility

A regularization term was included to discourage parameter estimates from moving unnecessarily far from their initial values. Let $\theta_i^{(0)}$ denote the initial value of parameter $i$, and let $u_i - l_i$ denote the width of its predefined calibration interval. The regularisation penalty was

$$E_{\text{reg}}(\boldsymbol{\theta}) = \frac{1}{17}\sum_{i=1}^{17}\left(\frac{\theta_i - \theta_i^{(0)}}{u_i - l_i}\right)^2.$$

To preserve the established excess risk of cervical cancer among women living with HIV, an additional soft penalty was applied to the cervical cancer incidence rate ratio between HIV-positive and HIV-negative women. A target value of 6 was used, informed by a large meta-analysis reporting a pooled relative risk of 6.07 (95% CI: 4.40–8.37) for cervical cancer among women living with HIV compared with women without HIV [2].

Age-specific incidence rate ratios were evaluated for age groups spanning 20–69 years. The penalty encouraged the modelled ratios to remain close to the target value of 6 while applying additional penalties to values below 1 or above 10. The interval 1–10 was used as a broad soft calibration constraint and should not be interpreted as an empirical confidence interval.

For a valid age group $g$, with incidence rate ratio $RR_g$, the contribution to the penalty was

$$\left[\log\left(\frac{RR_g}{6}\right)\right]^2 + \left[\frac{\max(1 - RR_g, 0)}{6}\right]^2 + \left[\frac{\max(RR_g - 10{,}0)}{6}\right]^2.$$

The overall rate-ratio penalty, $E_{\text{RR}}$, was defined as the mean of these contributions across valid age groups.

The complete objective function was therefore

$$E_{\text{total}}(\boldsymbol{\theta}) = E_{\text{data}}(\boldsymbol{\theta}) + 0.05E_{\text{reg}}(\boldsymbol{\theta}) + 0.05E_{\text{RR}}(\boldsymbol{\theta}).$$

Candidate parameter vectors that produced non-finite model outputs, non-positive Gaussian standard deviations, non-positive tunnel durations, or invalid model simulations were assigned a large objective value and were therefore excluded from the feasible solution space.

#### 2.4 Parameter bounds

All 17 calibrated parameters were restricted to predefined bounds (Appendix Table *A3*). The amplitude bounds for HPV acquisition and cervical cancer mortality were specified as annual probabilities in the calibration code and converted to continuous-time rates using $r = -\ln(1 - p)$. Bounds for the Gaussian location and dispersion parameters were expressed in years.

Table A3. Parameter bounds applied during country-specific model calibration.

| Parameter | Description | Lower bound | Upper bound |
|---|---|---|---|
| $A_{\text{HPV}}$ | Amplitude of the HPV acquisition hazard, HIV-negative | $-\ln(1-0.03)=0.0305$ | $-\ln(1-0.85)=1.8971$ |
| $m_{\text{HPV}}$ | Age at peak HPV acquisition, HIV-negative | 17 years | 45 years |
| $s_{\text{HPV}}$ | Dispersion of HPV acquisition by age, HIV-negative | 3 years | 25 years |
| $A^{+}_{\text{HPV}}$ | Amplitude of the HPV acquisition hazard, HIV-positive | $-\ln(1-0.05)=0.0513$ | $-\ln(1-0.60)=0.9163$ |
| $m^{+}_{\text{HPV}}$ | Age at peak HPV acquisition, HIV-positive | 18 years | 35 years |
| $s^{+}_{\text{HPV}}$ | Dispersion of HPV acquisition by age, HIV-positive | 4 years | 20 years |
| $A_{\mu_C}$ | Amplitude of cervical cancer mortality, HIV-negative | $-\ln(1-0.01)=0.0101$ | $-\ln(1-0.85)=1.8971$ |
| $m_{\mu_C}$ | Age at peak cervical cancer mortality, HIV-negative | 45 years | 90 years |
| $s_{\mu_C}$ | Dispersion of cervical cancer mortality by age, HIV-negative | 2.5 years | 30 years |
| $A^{+}_{\mu_C}$ | Amplitude of cervical cancer mortality, HIV-positive | $-\ln(1-0.01)=0.0101$ | $-\ln(1-0.90)=2.3026$ |
| $m^{+}_{\mu_C}$ | Age at peak cervical cancer mortality, HIV-positive | 35 years | 90 years |
| $s^{+}_{\mu_C}$ | Dispersion of cervical cancer mortality by age, HIV-positive | 3 years | 25 years |
| $A_{\text{HIV}}$ | Amplitude of the HIV acquisition hazard | $1\times10^{-6}$ | 0.0400 |
| $m_{\text{HIV}}$ | Age at peak HIV acquisition | 18 years | 45 years |
| $s_{\text{HIV}}$ | Dispersion of HIV acquisition by age | 3 years | 20 years |
| $T_P$ | Total duration of the three precancer states, HIV-negative | 18 years | 27 years |
| $T^{+}_P$ | Total duration of the three precancer states, HIV-positive | 12 years | 25 years |

Bounds applied to parameters estimated during model calibration

#### 2.5 Optimization

The objective function was minimized using the bound-constrained L-BFGS-B algorithm. The current analysis used one optimization start, initialized from the prespecified country parameter vector. The optimizer was allowed a maximum of 10,000 iterations and 150,000 objective-function evaluations. The calibrated parameter vector was the feasible solution with the smallest total objective value.

After calibration, model fit was assessed by comparing simulated and observed age-specific cervical cancer incidence, cervical cancer mortality, and HIV prevalence. The implied precancer durations, derived progression rates, incidence rate ratios, parameter-boundary positions, and population conservation were also examined as calibration diagnostics.

### Appendix 3. Health outcome and cost-effectiveness calculations

#### 3.1 Years of life lost (YLL)

Cervical cancer deaths were obtained from the annual differences in cumulative death registers for HIV-negative and HIV-positive women. Discounted years of life lost (YLL) were calculated using age-specific cervical cancer deaths and corresponding discounted remaining life expectancy:

$$YLL_{\text{disc}} = \sum_{a=0}^{100} \frac{d_{CC}(a)L_{\text{disc}}(a)}{(1+r)^a}$$

where $d_{CC}(a)$ represents new cervical cancer deaths at age $a$, $r$ is the annual health discount rate. Discounted remaining life expectancy was calculated as

$$L_{\text{disc}}(a) = \sum_{\tau=0}^{L(a)-1} \frac{1}{(1+r)^{\tau}}$$

where $L(a)$ represents the remaining life expectancy at age $a$ .

#### 3.2 Years lived with disability (YLD)

Years lived with disability (YLD) were calculated using a prevalence-based approach, considering the time spent living with invasive cervical cancer among HIV-negative and HIV-positive women. Cervical cancer person-years were estimated using the average number of women in the cancer compartments at the beginning and end of each annual cycle:

$$PY_{CC}(a) = \frac{C(a) + C(a+1)}{2} + \frac{C^{+}(a) + C^{+}(a+1)}{2}$$

where $C(a)$ and $C(a+1)$ represent the numbers of HIV-negative and HIV-positive women with cervical cancer at age $a$, respectively. Discounted YLD were calculated as

Discounted YLD were calculated as

$$YLD_{\text{disc}} = \sum_{a=0}^{99} \frac{PY_{CC}(a) \times DW_{CC}}{(1+r)^{a+0.5}}$$

where $DW_{CC}$ is the cervical cancer disability weight [8], and $r$ is the annual discount rate.

#### 3.3 Disability-adjusted life-years (DALYs)

Total discounted disability-adjusted life-years were calculated as

$$\text{DALYs}_{\text{disc}} = \text{YLL}_{\text{disc}} + \text{YLD}_{\text{disc}}.$$

For each vaccination strategy $s$, DALYs averted relative to the baseline strategy S0 were calculated as

$$\Delta E_s = \text{DALYs}_{\text{disc}}(\text{S0}) - \text{DALYs}_{\text{disc}}(s).$$

#### 3.4 Vaccination programme costs

Vaccination programme costs were calculated based on the number of physical vaccine doses administered. Routine vaccination consisted of two doses, primary catch-up vaccination among previously unvaccinated women living with HIV consisted of three doses, and booster vaccination consisted of one dose.

The total number of physical vaccine doses administered under strategy $s$ at age $a$ was calculated as

$$N_{\text{dose},s}(a) = 2N_{\text{routine},s}(a) + 3N_{\text{catchup},s}(a) + N_{\text{boost},s}(a).$$

where $N_{\text{routine},s}(a)$, $N_{\text{catchup},s}(a)$, and $N_{\text{boost},s}(a)$ represent the numbers of women receiving routine, catch-up, and booster vaccination, respectively.

Discounted vaccination programme costs were calculated as

$$C_{\text{vacc},s} = \sum_{a=0}^{100} \frac{c_{\text{dose}}\, N_{\text{dose},s}(a)}{(1+r)^{a}}$$

where $c_{\text{dose}}$ is the cost per physical vaccine dose, including vaccine acquisition and delivery, and $r$ is the annual discount rate.

#### 3.5 Country-specific cervical cancer treatment unit costs

Country-specific cervical cancer treatment costs were estimated using Tanzania as the reference country. For each country \(i\), the unit cost per incident cervical cancer case was calculated as

$$c_{CC,i} = c_{CC,\text{Tanzania}} \times \frac{GDPpc_i}{GDPpc_{\text{Tanzania}}}$$

where $C_{CC,\text{Tanzania}} = 2{,}526$ USD is the reference treatment cost per case, and $GDPpc_i$ and $GDPpc_{\text{Tanzania}}$ represent the GDP per capita of country $i$ and Tanzania, respectively. GDP per capita values were expressed in current US dollars for 2025.

#### 3.6 Cervical cancer treatment costs

Cervical cancer treatment costs were calculated using the number of new cervical cancer cases and the country-specific treatment cost per case. Discounted treatment costs under strategy \(s\) were calculated as cancer tre

$$C_{CC,s} = \sum_{a=0}^{100} \frac{c_{CC} \times Cases_s(a)}{(1+r)^a}$$

where $Cases_s(a)$ represents new cervical cancer cases at age $a$, summed across HIV-negative and HIV-positive women, $c_{CC}$ is the country-specific treatment cost per case, and $r$ is the annual discount rate.

Under the health-system perspective, total costs were

$$C_{\text{health},s} = C_{\text{vacc},s} + C_{\text{CC},s}.$$

#### 3.7 Incremental costs and avoided treatment expenditure

Incremental costs were calculated as the difference in total costs between each vaccination $s$ and the baseline strategy S0:

$$\Delta C_s = C_s - C_{\text{S0}}.$$

Under the health-system perspective, avoided cervical cancer treatment costs were calculated as

$$C_{\text{avoided},s} = C_{\text{CC,S0}} - C_{\text{CC},s}.$$

The net incremental health-system cost can therefore also be written as

$$\Delta C_{\text{health},s} = \Delta C_{\text{vacc},s} - C_{\text{avoided},s},$$

Where $\Delta C_{\text{vacc},s}$ represents the additional vaccination programme cost of strategy $s$ relative to S0.

#### 3.8 Direct incremental cost-effectiveness ratios (ICERs)

The incremental cost-effectiveness ratio (ICER) for each vaccination strategy $s$ relative to S0 was calculated as

$$\text{ICER}_s = \frac{\Delta C_s}{\Delta E_s}$$

where $\Delta C_s$ represents the incremental cost and $\Delta E_s$ represents DALYs averted relative to S0.

Strategies with negative incremental costs ( $\Delta C_s < 0$ ) and positive health gains ($\Delta E_s > 0$)

were classified as cost-saving.

#### 3.9 Sequential cost-effectiveness analysis

Strategies were ranked by increasing effectiveness, excluding strictly and extendedly dominated strategies. Sequential ICERs were calculated between consecutive strategies on the cost-effectiveness frontier as

$$\text{ICER}_j^{\text{seq}} = \frac{C_j - C_{j-1}}{E_j - E_{j-1}},$$

where $C_j$ represents the total cost $E_j$ denotes DALYs averted relative to S0.
The sequential ICER represents the additional cost per additional DALY averted when moving to the next more effective str ategy.

#### 3.10 Marginal net cost per additional vaccine dose

The net incremental health-system cost was standardised per 100,000 additional physical vaccine doses relative to S0:

$$M_s = \frac{\Delta C_{\text{health},s}}{N_{\text{dose},s} - N_{\text{dose,S0}}} \times 100{,}000.$$

where $\Delta C_{\text{health},s}$ represents the incremental health-system cost and $N_{\text{dose},s}$ represents the total number of physical vaccine doses under strategy $s$.

Negative values indicate net cost savings, while positive values indicate additional expenditure.

### Appendix 4. Sensitivity analyses

#### 4.1 Deterministic one-way sensitivity analysis

A deterministic one-way sensitivity analysis (OWSA) was conducted for Mozambique to examine the effect of uncertainty in calibrated epidemiological parameters on model outcomes. The analysis followed a one-at-a-time design: each parameter was varied independently while the remaining parameters were fixed at their calibrated values.

The analysis included 17 calibrated parameters: the amplitude, peak age, and spread of five age-dependent functions describing HPV acquisition among HIV-negative and HIV-positive women, cervical cancer mortality in both HIV strata, and HIV acquisition (15 parameters); and the duration of precancerous disease in each HIV stratum (two parameters). Each parameter was varied across five equally spaced values from 80% to 120% of its calibrated value (0.80, 0.90, 1.00, 1.10, and 1.20). This yielded 85 parameter settings (17 parameters × 5 values). When precancer duration was varied, its associated progression rates were recalculated.

For each setting, the model was run under the baseline vaccination strategy (S0). The outcomes were (i) total discounted DALYs, calculated using the revised prevalence-based disability component, (ii) total discounted health-system costs, and (iii) total physical vaccine doses administered. For each parameter, the minimum and maximum changes in each outcome relative to the calibrated S0 result were retained. Local sensitivity was evaluated using a central finite-difference approximation around the calibrated parameter value:

$$\frac{\partial Y}{\partial p_j} \approx \frac{Y\left(1.10\hat{p}_j\right) - Y\left(0.90\hat{p}_j\right)}{0.20\hat{p}_j},$$

where $Y$ denotes an S0 model outcome and $\hat{p}_j$ is the calibrated value of parameter $j$. Results were presented in paired tornado diagrams showing the min–max changes and the signed local derivatives. Parameter settings producing invalid model outputs were excluded.

#### 4.2 Probabilistic sensitivity analysis

A probabilistic sensitivity analysis (PSA) was conducted to evaluate the joint effect of uncertainty in the 17 calibrated parameters. In each of 2,000 iterations, all parameters were sampled simultaneously from independent uniform distributions within ±10% of their calibrated values:

$$p_j^{(k)} \sim \mathcal{U}\left(0.90\hat{p}_j,\ 1.10\hat{p}_j\right).$$

The same valid sampled parameter vector was used to simulate the baseline strategy (S0) and all intervention strategies (S1–S6). For each strategy $s$, incremental health-system cost was calculated as $\Delta C_s = C_s - C_{S0}$, and discounted DALYs averted as $\Delta E_s = DALY_{S0} - DALY_s$. The resulting distributions were summarised using medians, 2.5th–97.5th percentile uncertainty intervals, and cost-effectiveness planes.

### Appendix 5. Supplementary inputs, calibration and economic results

#### 5.1 Epidemiological and demographic input data

To ensure consistency across countries, epidemiological and demographic inputs were obtained from the Global Burden of Disease study for the 2023 reference year [26]. The analysis included 13 countries: Cameroon, Côte d'Ivoire, Gambia, Kenya, Lesotho, Malawi, Mozambique, Rwanda, Tanzania, Togo, Uganda, Zambia, and Zimbabwe. All inputs were restricted to females and extracted using consistent age groups, outcome definitions, and measures across countries.

Table A4. Epidemiological and demographic inputs used in model calibration.

| Input | Measure | Use in the model |
|---|---|---|
| Cervical cancer incidence | Age-specific rate per 100,000 females | Calibration of cervical cancer incidence |
| Cervical cancer mortality | Age-specific rate per 100,000 females | Calibration of cervical cancer mortality |
| HIV prevalence | Age-specific percentage among females | Calibration of the HIV acquisition hazard |
| Background mortality | Age-specific probability of death from the female life table | Background mortality from causes other than HIV and cervical cancer |
| HIV-related mortality | Age-specific probability of death attributed to HIV/AIDS | Additional mortality among HIV-positive individuals |
| Remaining life expectancy | Expected remaining years of life by age | Calculation of years of life lost following cervical cancer death |

Epidemiological and demographic inputs obtained from the Global Burden of Disease study [26]

The calibration targets comprised age-specific cervical cancer incidence, cervical cancer mortality, and HIV prevalence. Cervical cancer incidence and mortality were expressed as rates per 100,000 women, while HIV prevalence was expressed as a percentage of the female population. Model outputs were aggregated to the same age groups as the empirical data before comparison during calibration.

Age-specific background mortality was derived from female life-table probabilities of death. HIV-related mortality was obtained from cause-specific mortality estimates attributed to HIV/AIDS. Remaining female life expectancy at each age was used to calculate years of life lost following cervical cancer death. The use of a common data source and reference year improved consistency and comparability across countries.

The country-specific calibration targets and corresponding model predictions are presented in Figure A1.

#### 5.2 Country-specific model calibration

The model was calibrated separately for each country using age-specific cervical cancer incidence, cervical cancer mortality, and HIV prevalence data. Model outputs were aggregated to the same age groups as the observed data and compared during optimization. The resulting country-specific fits are shown in Figure A1, where model predictions are presented alongside the corresponding observed values.

Table A5. Country-specific calibrated epidemiological parameters for the 13 study countries.

| Parameter | Cameroon | Côte d'Ivoire | Gambia | Kenya | Lesotho | Malawi | Mozambique | Rwanda | Tanzania | Togo | Uganda | Zambia | Zimbabwe |
|---|---|---|---|---|---|---|---|---|---|---|---|---|---|
| **HPV incidence, HIV-negative** | | | | | | | | | | | | | |
| **Amplitude** | 1.6402 | 1.8971 | 0.8664 | 1.8971 | 1.8971 | 1.8971 | 0.7584 | 1.8971 | 1.5814 | 1.8971 | 1.8971 | 1.4064 | 1.8971 |
| **Peak age** | 45.00 | 45.00 | 29.47 | 43.14 | 45.00 | 39.33 | 45.00 | 19.60 | 45.00 | 45.00 | 41.29 | 24.84 | 45.00 |
| **Spread** | 17.73 | 23.30 | 25.00 | 25.00 | 18.78 | 25.00 | 21.93 | 25.00 | 14.37 | 25.00 | 25.00 | 7.08 | 25.00 |
| **HPV incidence, HIV-positive** | | | | | | | | | | | | | |
| **Amplitude** | 0.3455 | 0.5070 | 0.5531 | 0.5478 | 0.2268 | 0.9163 | 0.0562 | 0.9163 | 0.9163 | 0.4541 | 0.0756 | 0.2453 | 0.1026 |
| **Peak age** | 20.83 | 25.01 | 20.91 | 19.47 | 35.00 | 21.83 | 18.81 | 24.82 | 35.00 | 18.29 | 19.51 | 35.00 | 19.92 |
| **Spread** | 10.11 | 4.00 | 4.00 | 4.00 | 20.00 | 7.62 | 19.45 | 7.90 | 13.09 | 4.01 | 12.61 | 20.00 | 20.00 |
| **Cervical cancer mortality, HIV-negative** | | | | | | | | | | | | | |
| **Amplitude** | 1.7185 | 0.8787 | 0.9395 | 0.8074 | 1.8971 | 0.3578 | 1.8971 | 0.4925 | 1.7661 | 0.9331 | 0.6294 | 0.9103 | 1.8971 |
| **Peak age** | 81.29 | 81.36 | 89.72 | 86.39 | 81.70 | 82.34 | 77.94 | 90.00 | 90.00 | 89.63 | 86.01 | 87.69 | 80.17 |
| **Spread** | 21.65 | 24.29 | 29.99 | 11.15 | 25.81 | 10.81 | 20.35 | 15.22 | 12.26 | 27.66 | 11.91 | 7.05 | 21.74 |
| **Cervical cancer mortality, HIV-positive** | | | | | | | | | | | | | |
| **Amplitude** | 1.6344 | 0.1147 | 1.1689 | 0.2852 | 0.2839 | 0.3259 | 0.0406 | 2.3026 | 1.8926 | 0.1596 | 0.1891 | 1.7764 | 0.1807 |
| **Peak age** | 89.91 | 71.72 | 77.77 | 44.43 | 90.00 | 35.00 | 61.24 | 52.70 | 79.40 | 89.94 | 46.79 | 80.32 | 90.00 |
| **Spread** | 7.60 | 19.42 | 3.23 | 25.00 | 25.00 | 25.00 | 24.95 | 18.98 | 17.47 | 24.87 | 25.00 | 25.00 | 24.37 |
| **HIV incidence** | | | | | | | | | | | | | |
| **Amplitude** | 0.00722 | 0.00666 | 0.00463 | 0.00959 | 0.04000 | 0.01963 | 0.02623 | 0.00595 | 0.00973 | 0.00506 | 0.01305 | 0.02246 | 0.02334 |
| **Peak age** | 32.70 | 33.52 | 30.16 | 31.01 | 26.82 | 31.14 | 23.17 | 34.90 | 28.40 | 32.46 | 28.10 | 28.77 | 30.13 |
| **Spread** | 9.39 | 8.97 | 8.09 | 8.33 | 8.76 | 8.22 | 5.55 | 8.44 | 7.64 | 7.55 | 7.07 | 7.35 | 7.55 |
| **Precancer duration** | | | | | | | | | | | | | |
| **HIV-negative** | 22.87 | 24.31 | 24.65 | 24.02 | 22.86 | 21.14 | 22.98 | 22.04 | 20.66 | 23.69 | 23.17 | 19.19 | 25.35 |
| **HIV-positive** | 17.09 | 16.16 | 16.38 | 13.92 | 21.29 | 12.00 | 21.02 | 12.00 | 15.48 | 14.32 | 12.00 | 14.82 | 24.81 |

Five age-dependent functions are characterized by amplitude, peak age and spread. Precancer durations indicate total time across the three sequential precancer states.

Note: Peak age and spread are in years. Calibrated values were extracted from the final country-specific 17-parameter vectors. HPV incidence, cervical cancer mortality and HIV incidence each have Gaussian amplitude, peak-age and spread parameters.

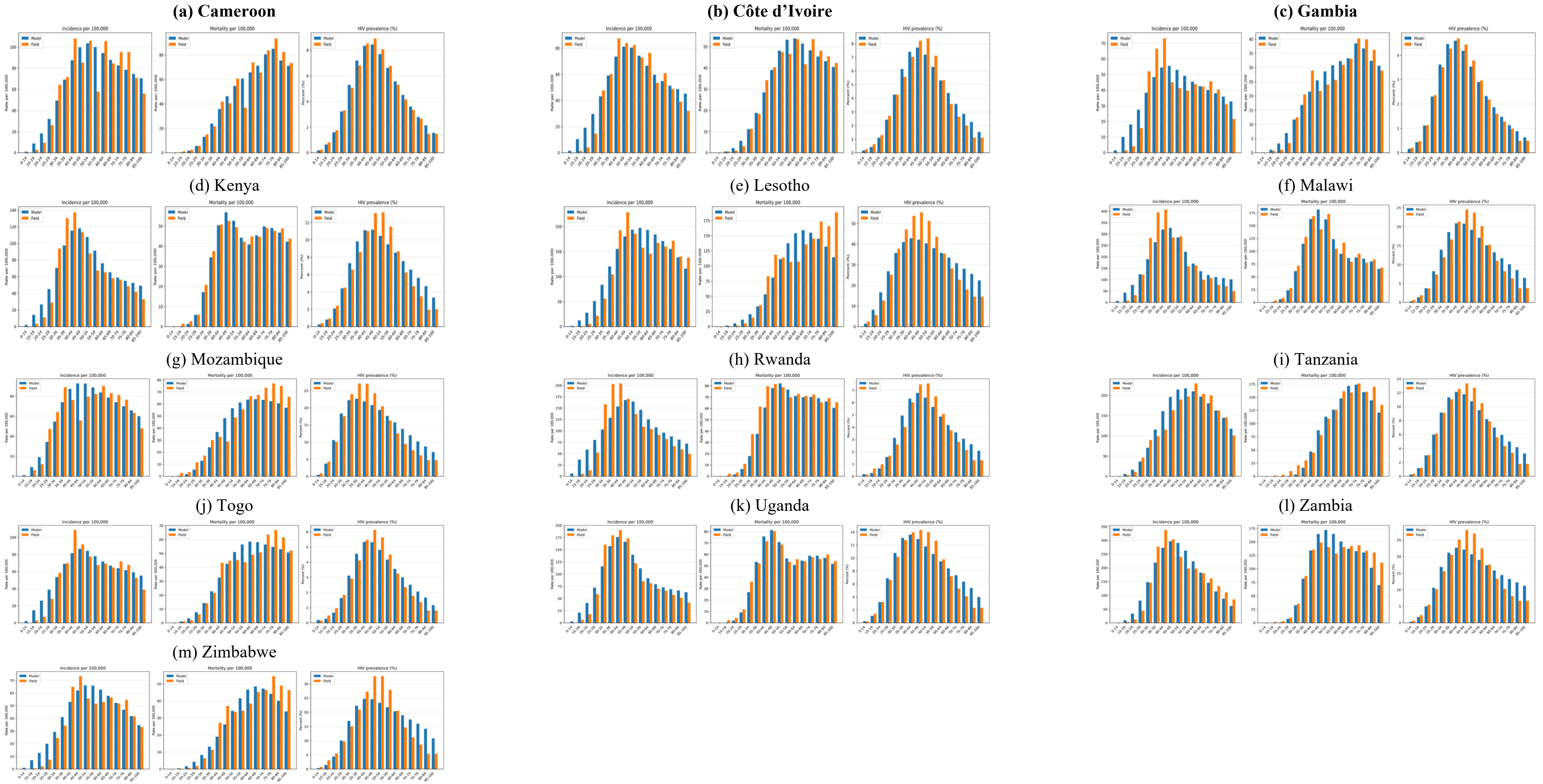


Figure A1. Comparison of model predictions and observed age-specific cervical cancer incidence, cervical cancer mortality, and HIV prevalence across 13 countries. Blue bars indicate model predictions; orange bars indicate observed data.

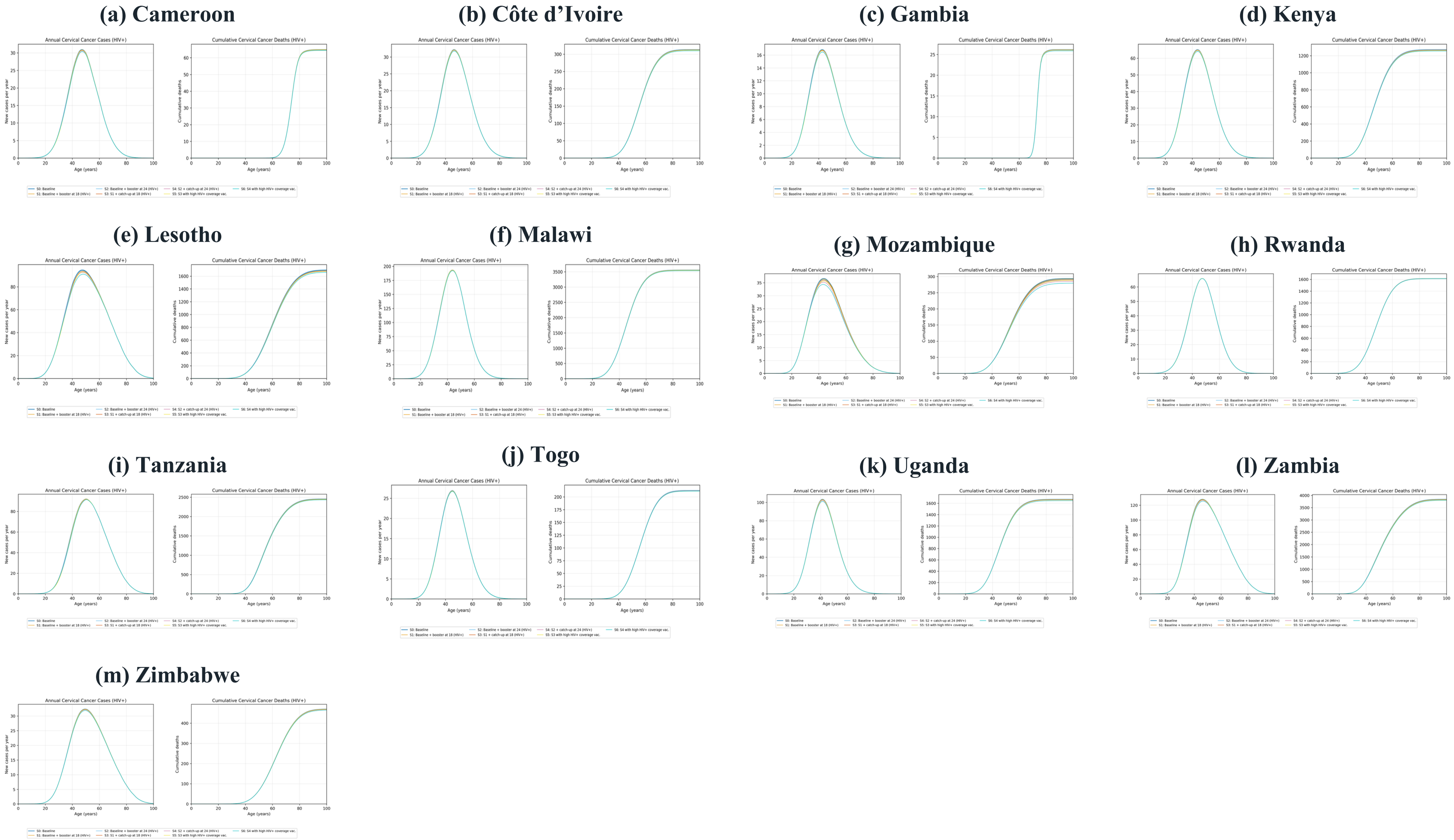


Figure A2. Annual cervical cancer cases and cumulative cervical cancer deaths among women living with HIV across the 13 study countries under vaccination strategies S0–S6.

#### 5.3 Additional cost-effectiveness results

Table A6. Sequential ICERs of HIV-targeted HPV vaccination strategies across 13 African countries, including cervical cancer management costs.

| Strategy | Cameroon | Côte d’Ivoire | Gambia | Kenya | Lesotho | Malawi | Mozambique | Rwanda | Tanzania | Togo | Uganda | Zambia | Zimbabwe |
|---|---|---|---|---|---|---|---|---|---|---|---|---|---|
| **S0. Baseline routine vaccination** | – | – | – | – | – | – | – | – | – | – | – | – | – |
| **S1. Baseline + booster at age 18 (HIV+)** | – | – | – | – | – | – | – | – | – | – | – | – | – |
| **S2. Baseline + booster at age 24 (HIV+)** | – | – | – | – | – | – | – | – | – | – | – | – | – |
| **S3. S1 + three-dose catch-up primary at age 18 (HIV+), standard coverage** | – | – | – | – | 39.85 | – | 491.96 | – | – | – | – | – | – |
| **S4. S2 + three-dose catch-up primary at age 24 (HIV+), standard coverage** | – | – | – | – | – | – | 646.42 | – | – | – | – | – | – |
| **S5. S3 with high HIV+ coverage (98%)** | – | – | – | – | 102.26 | – | 534.76 | – | – | – | – | – | – |
| **S6. S4 with high HIV+ coverage (98%)** | – | – | 1,033.69 | – | 278.05 | – | 678.70 | – | – | – | – | – | 279.69 |

Note: Sequential ICERs are in USD per additional discounted DALY averted. Dash indicates that no sequential ICER applies (the strategy is dominated or is the first strategy on the frontier).

Table A7. Sequential ICERs of HIV-targeted HPV vaccination strategies across 13 African countries, excluding cervical cancer management costs.

| Strategy | Cameroon | Côte d’Ivoire | Gambia | Kenya | Lesotho | Malawi | Mozambique | Rwanda | Tanzania | Togo | Uganda | Zambia | Zimbabwe |
|---|---|---|---|---|---|---|---|---|---|---|---|---|---|
| **S0. Baseline routine vaccination** | – | – | – | – | – | – | – | – | – | – | – | – | – |
| **S1. Baseline + booster at age 18 (HIV+)** | 204.41 | 67.76 | 144.70 | 10.70 | 204.82 | 10.28 | – | 8.44 | – | 41.42 | 20.74 | – | – |
| **S2. Baseline + booster at age 24 (HIV+)** | 135.49 | 35.77 | – | – | 76.99 | 7.89 | 189.88 | 6.35 | 15.15 | – | – | 15.02 | 166.42 |
| **S3. S1 + three-dose catch-up primary at age 18 (HIV+), standard coverage** | 347.05 | – | 375.32 | 28.82 | 335.38 | 31.55 | 668.76 | 28.46 | 36.57 | 95.09 | 72.17 | 52.33 | 725.73 |
| **S4. S2 + three-dose catch-up primary at age 24 (HIV+), standard coverage** | – | – | – | – | – | – | – | – | – | – | – | – | – |
| **S5. S3 with high HIV+ coverage (98%)** | 531.26 | – | 510.97 | 37.25 | 394.19 | 39.39 | 729.94 | 31.96 | 70.08 | 132.57 | 77.85 | 72.66 | 766.13 |
| **S6. S4 with high HIV+ coverage (98%)** | 1,010.63 | 176.64 | 1,742.90 | 277.97 | 511.51 | 62.77 | 881.62 | 72.46 | 79.14 | – | 138.01 | 96.15 | 1,045.88 |

Note: Sequential ICERs are in USD per additional discounted DALY averted. Dash indicates that no sequential ICER applies (the strategy is dominated or is the first strategy on the frontier).

#### 5.4 HPV vaccination assumptions

Table A8. HPV vaccination assumptions used in the base-case analysis, including vaccination schedule, physical doses, vaccine efficacy and duration of protection.

| Population / target group | Vaccination schedule in the model | Physical doses | Assumed initial efficacy | Assumed waning period | Evidence and modelling assumptions |
|---|---|---|---|---|---|
| HIV-negative girls eligible for routine vaccination | Routine primary vaccination at age 14 | 2 | 96% | 20 years | Evidence supports high efficacy and sustained protection following HPV vaccination in immunocompetent populations [5, 4]. The initial efficacy and linear waning period were specified as modelling assumptions. |
| HIV-positive girls eligible for routine vaccination | Routine primary vaccination at age 14 | 2 | 87% | 10 years | HPV vaccines have demonstrated favourable safety and immunogenicity among people living with HIV [6, 15]. Lower initial efficacy and a shorter waning period were assumed to reflect uncertainty regarding long-term clinical protection. |
| Previously unvaccinated women living with HIV targeted by S3–S6 | Primary catch-up vaccination at age 18 or 24 | 3 | 87% | 10 years | WHO recommends at least two doses and, where feasible, three doses for people living with HIV [4]. The same efficacy and waning assumptions as routine vaccination among HIV-positive individuals were applied, reflecting limited evidence on long-term protection [6, 15]. |
| Previously vaccinated women living with HIV targeted by S1–S6 | One additional booster dose at age 18 or 24 | 1 additional dose | 96% | 30 years following boosting | Booster vaccination was assumed to restore protection and reset the waning period. Given limited evidence on long-term clinical booster effectiveness among women living with HIV, these parameters were treated as exploratory modelling assumptions [6, 27, 15]. |

### 5.5 Country-specific cervical cancer management costs

Because comparable empirical cervical cancer management costs were not available for all study countries, country-specific unit costs were extrapolated from the Tanzanian reference value using relative GDP per capita. The Tanzania reference cost was USD 2,526 per incident case [11]. GDP per capita values were measured in current US dollars for 2025 [12]

Table A9. Country-specific cervical cancer management cost inputs.

| Country | GDP per capita (current USD, 2025) | GDP ratio vs Tanzania | Estimated cancer cost (USD per case) |
|---|---:|---:|---:|
| Cameroon | 1,972.40 | 1.496 | 3,777.89 |
| Côte d'Ivoire | 3,050.10 | 2.313 | 5,842.09 |
| Gambia | 919.10 | 0.697 | 1,760.42 |
| Kenya | 2,362.90 | 1.792 | 4,525.85 |
| Lesotho | 1,089.00 | 0.826 | 2,085.85 |
| Malawi | 671.50 | 0.509 | 1,286.18 |
| Mozambique | 626.90 | 0.475 | 1,200.75 |
| Rwanda | 1,123.70 | 0.852 | 2,152.31 |
| Tanzania | 1,318.80 | 1.000 | 2,526.00 |
| Togo | 1,383.90 | 1.049 | 2,650.69 |
| Uganda | 1,206.30 | 0.915 | 2,310.52 |
| Zambia | 1,317.90 | 0.999 | 2,524.28 |
| Zimbabwe | 3,021.40 | 2.291 | 5,787.12 |

Notes: GDP per capita values are expressed in current US dollars for 2025. The GDP ratio was calculated as the GDP per capita of each country divided by the GDP per capita of Tanzania. Estimated cancer cost was calculated as $2{,}526 \times (GDPpc_i / GDPpc_{Tanzania})$. The Tanzania estimate was used as reported in the source study and was not separately adjusted for inflation. The extrapolated costs are modelling inputs and do not represent empirical costing studies in the respective countries. Costs for countries other than Tanzania were extrapolated from the Tanzanian reference value of USD 2,526 using the ratio of country GDP per capita to Tanzanian GDP per capita.

### 5.6 Supplementary sensitivity results

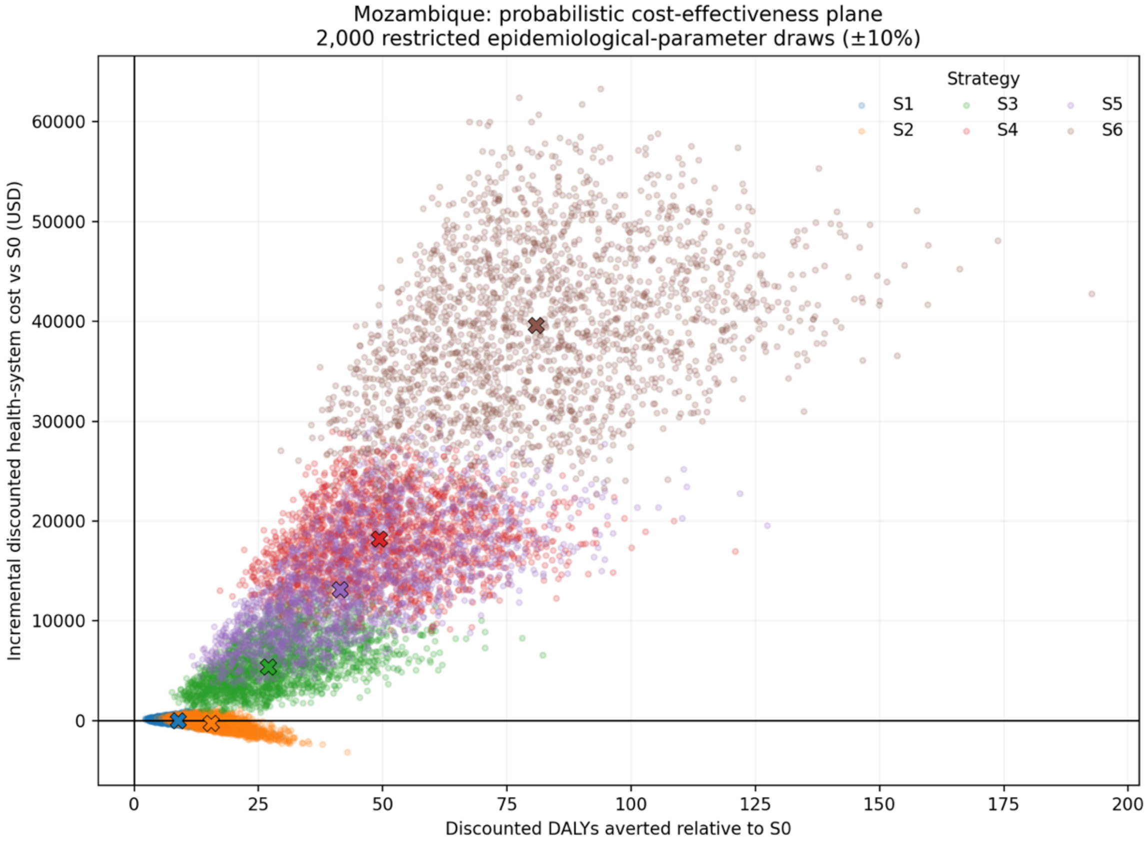


Figure A3. Probabilistic cost-effectiveness plane for Mozambique based on 2,000 sampled sets of the 17 calibrated epidemiological parameters (±10%). Each point indicates incremental discounted DALYs averted and incremental health-system cost relative to S0 for one vaccination strategy in one simulation.

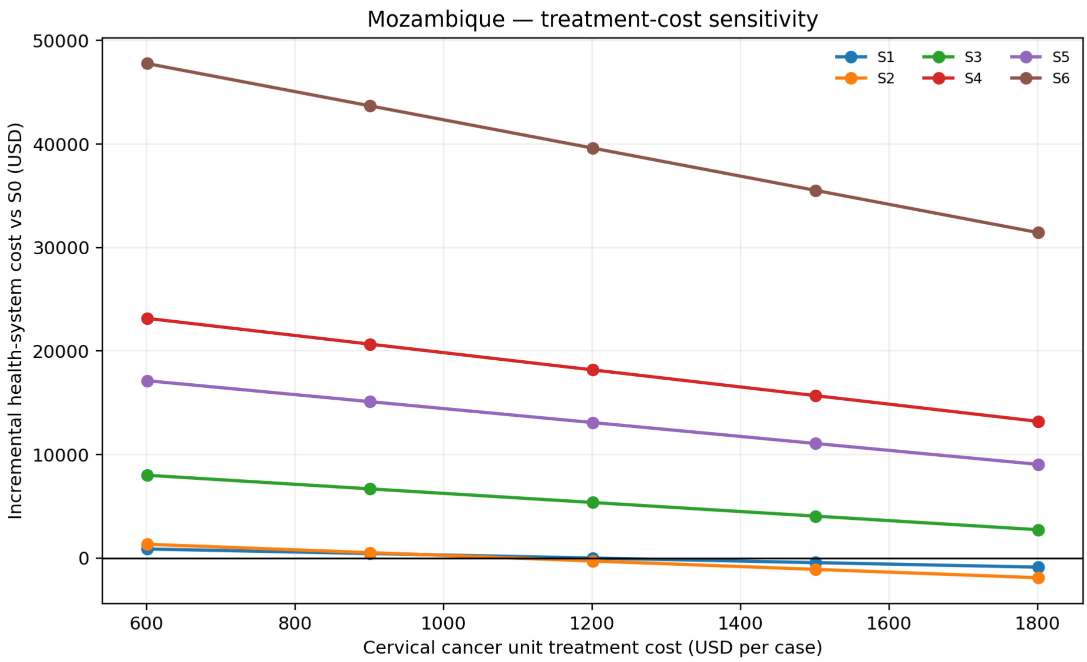


Figure A4. Sensitivity of incremental health-system costs to the cervical cancer unit treatment cost in Mozambique, under vaccination strategies S1–S6 relative to S0. Negative incremental costs indicate net cost savings.
